# Systematic review of the scrub typhus treatment landscape: Assessing the feasibility of an individual participant-level data (IPD) platform

**DOI:** 10.1101/2021.03.31.21254664

**Authors:** Kartika Saraswati, Brittany J Maguire, Alistair RD McLean, Sauman Singh-Phulgenda, Roland C Ngu, Paul N Newton, Nicholas PJ Day, Philippe J Guérin

## Abstract

**Background:** Scrub typhus is an acute febrile illness caused by intracellular bacteria from the genus *Orientia*. It is estimated that one billion people are at risk, with one million cases annually mainly affecting rural areas in Asia-Oceania. Relative to its burden, scrub typhus is understudied, and treatment recommendations vary with poor evidence base. These knowledge gaps could be addressed by establishing an individual participant-level data (IPD) platform, which would enable pooled, more detailed and statistically powered analyses to be conducted. This study aims to assess the characteristics of scrub typhus treatment studies and explore the feasibility and potential value of developing a scrub typhus IPD platform to address unanswered research questions.

**Methodology/Principal Findings:** We conducted a systematic literature review looking for prospective scrub typhus clinical treatment studies published from 1998 to 2020. Six electronic databases (Ovid Embase, Ovid Medline, Ovid Global Health, Cochrane Library, Scopus, Global Index Medicus), ClinicalTrials.gov, and WHO ICTRP were searched. We extracted data on study design, treatment tested, patient characteristics, diagnostic methods, geographical location, outcome measures, and statistical methodology.

Among 3,100 articles screened, 127 were included in the analysis. 12,079 participants from 12 countries were enrolled in the identified studies. ELISA, PCR, and eschar presence were the most commonly used diagnostic methods. Doxycycline, azithromycin, and chloramphenicol were the most commonly administered antibiotics. Mortality, complications, adverse events, and clinical response were assessed in most studies. There was substantial heterogeneity in the diagnostic methods used, treatment administered (including dosing and duration), and outcome assessed across studies. There were few interventional studies and limited data collected on specific groups such as children and pregnant women.

**Conclusions/Significance:** There were a limited number of interventional trials, highlighting that scrub typhus remains a neglected disease. The heterogeneous nature of the available data reflects the absence of consensus in treatment and research methodologies and poses a significant barrier to aggregating information across available published data without access to the underlying IPD. There is likely to be a substantial amount of data available to address knowledge gaps. Therefore, there is value for an IPD platform that will facilitate pooling and harmonisation of currently scattered data and enable in-depth investigation of priority research questions that can, ultimately, inform clinical practice and improve health outcomes for scrub typhus patients.

**Author summary:** Scrub typhus is a febrile illness most commonly found in rural tropical areas. It is caused by a Gram-\negative bacteria belonging to the family Rickettsiaceae and transmitted by mites when they feed on vertebrates. There is an estimate of one million cases annually, with an estimated one billion people at risk, mostly in Asia-Oceania. But relative to the scale of the problem, scrub typhus is largely understudied. Evidence-based treatment recommendations by policymakers vary or are non-existent. We searched databases and registries for prospective scrub typhus clinical treatment studies published from 1998 to 2020 and reviewed them. Data from clinical trials and particularly for specific groups, such as pregnant women and children, were minimal. The methods used to measure treatment efficacy were heterogeneous, making it difficult to directly compare or conduct a meta-analysis based on aggregated data. One way to improve the current level of evidence would be by pooling and analysing individual participant-level data (IPD), i.e. the raw data from individual participants in completed studies. This review demonstrated that there is scope for developing a database for individual participant data to enable more detailed analyses. IPD meta-analyses could be a way to address knowledge gaps such as optimum dosing for children.

## Introduction

Scrub typhus is a zoonotic disease transmitted during feeding by larval mites from the genus *Leptotrombidium* [1]. It is caused by obligate intracellular bacteria from the genus *Orientia* [1]. *Orientia tsutsugamushi*, the main causal species, is prevalent within the so-called “tsutsugamushi triangle” – an area bordering Pakistan in the west, Far-East Russia in the northeast, and North Australia in the south [1]. Other novel species have also been detected outside the traditional tsutsugamushi triangle, i.e. *Candidatus* Orientia chuto and *Candidatus* Orientia chiloensis in Dubai and Chile, respectively [2, 3]. Scrub typhus largely affects rural populations for whom access to health care facilities and laboratories with diagnostic capacity are limited [1]. There are estimates of one billion people at risk and one million cases annually with the majority of the population at risk residing in low and middle income countries (LMICs) [4].

Scrub typhus manifests as an acute febrile illness. The median estimated mortality, if treated, is 1.4% (range 0-33%) [4]. If untreated, the median estimated mortality is 6% (range 0-70%) [5]. Typical clinical manifestations include fever, headache, myalgia, rash, and lymphadenopathy [1]. It is difficult to diagnose clinically because it lacks reliable distinguishing features that differentiate it from other febrile illnesses prevalent in the endemic areas, such as typhoid fever, dengue fever and malaria [1, 6]. Therefore, laboratory tests play an essential role in diagnosis. The gold standard for diagnosis is the indirect immunofluorescence assay (IFA), which requires sophisticated laboratory facilities and skilled personnel [6]. Antibiotics, including doxycycline, tetracycline, chloramphenicol, and azithromycin, are the current mainstays of treatment [1, 7]. The treatment efficacy of other antibiotics such as rifampicin has also been studied, although using it in areas with a high prevalence of tuberculosis is not recommended [8]. Delayed response to antibiotic treatment has been reported [9], although the contribution of antimicrobial resistance is uncertain [10]. National treatment recommendations vary or are simply not stated, and the absence of WHO recommendations illustrates the lack of high-quality supportive evidence for optimal treatments [7, 11].

The difficulty in diagnosis is one of the main reasons for the limited epidemiological knowledge on scrub typhus. The IFA has been used to diagnose scrub typhus since at least the 1960s [12], but although considered the gold standard, its accuracy is limited [13]. With scant and scattered data, it is difficult to identify the most at-risk population, the regional distribution, and the optimal treatments to optimise disease control efforts, which to date rely on limited evidence. Despite its mortality and morbidity impact, scrub typhus is not considered in the current WHO list of Neglected Tropical Diseases (NTD).

Existing data collected from clinical trials and longitudinal patient observational studies are the sources of our current evidence-based knowledge. However, there have been no recent reviews of the existing literature, and we hypothesise that in the context of scarce data, they are an underutilised source of information to address research priorities and bridge knowledge gaps. This systematic review aims to assess the characteristics of scrub typhus treatment studies and explore the need for and the feasibility of answering knowledge gaps through pooling scrub typhus individual patient data.

## Methods

### Search strategy

We conducted a systematic literature review following the Preferred Reporting Items for Systematic Reviews and Meta-Analyses (PRISMA) statement (S1 Checklist)[14]. The review is registered in the International Prospective Register of Systematic Reviews (PROSPERO) under the reference CRD42018089405. Six electronic databases (Ovid Embase, Ovid Medline, Ovid Global Health, Cochrane Library, Scopus, Global Index Medicus), ClinicalTrials.gov, and WHO ICTRP were searched using a search strategy detailed in the supporting information (S2 Appendix). The search was limited to publication dates from January 1998 up to February 2020 due to the significantly reduced likelihood of gathering individual participant-level data with time since publication [15]. No language restriction was applied at the search stage or initial screening of literature for eligibility. The reference lists of systematic reviews and meta-analyses (including Cochrane reviews) identified during the systematic search were screened for any additional studies that met the inclusion criteria to ensure comprehensive inclusion of articles.

### Eligibility criteria and study selection

Prospective clinical studies in humans with scrub typhus where treatment was administered and participants were followed up for at least 24 hours (i.e. clinical trials and longitudinal observational studies) were eligible for inclusion.

The exclusion criteria were as follows: studies on scrub typhus published earlier than 1 January 1998, conference abstracts published earlier than 1 January 2016, secondary research, case report/series, studies including five patients or less, studies on co-infection cases only, studies using non-definitive diagnosis i.e. not using confirmatory laboratory test, studies not yet completed, and studies without an accessible full-text. Although the search strategy and title/abstract screening were not restricted by language, due to the review team’s proficiencies, potentially relevant studies identified in languages other than English, French and Spanish were not included for data extraction and further analysis.

Titles and abstracts were screened by one researcher (KS). Full texts were independently reviewed by two authors (KS and SS) with Covidence systematic review software (Veritas Health Innovation, Melbourne, Australia). When conflicts arose, a third researcher (BM) was consulted. Inclusion was not limited to any minimum quality assessment to ensure a complete assessment of the research landscape.

### Data extraction and analysis

Relevant information from included studies was extracted using a piloted REDCap (Research Electronic Data Capture) database [16]. Extracted data included details on study design, treatment tested, patient characteristics, diagnostic methods, geographical location, outcome measures, and statistical methodology. Data extraction was performed by one of three reviewers (KS or SS or RN). For each publication, a second alternative reviewer (KS or SS or RN) cross-checked every data variable entered by the first reviewer for quality control to identify missed or discrepant information. The third reviewer resolved any data interpretation differences that arose between the two reviewers.

For the purposes of this review, we use the term “study arm” to refer both to a study arm from a clinical trial and patient cohorts from observational studies where treatment is administered.

Analysis was done with R version 3.6.2 and R Studio Version 1.2.5033 [17, 18]. The extracted data were synthesised using descriptive statistics, graphs and narrative synthesis. Categorical variables were summarised with proportions and frequencies. Continuous variables were summarised as quartiles, minimums and maximums.

## Results

### Search results

Formal database searches were first conducted on 23 March 2018, with a subsequent update search completed up to 17 February 2020, which identified a total of 6,134 citations. After removing duplicates across the primary and update searches, we screened the title and abstract of 3,100 articles (Fig 1). There were 660 full texts assessed for eligibility. From these, 533 articles were excluded, among those 357 were case reports or case series. We found 20 papers not in English: four in Korean and 16 in Chinese. We could not access the full text of six papers despite extensive attempts. We included 127 relevant studies in the analysis. There were seven interventional studies and 120 observational studies included (S3 Appendix).

**Fig 1.**
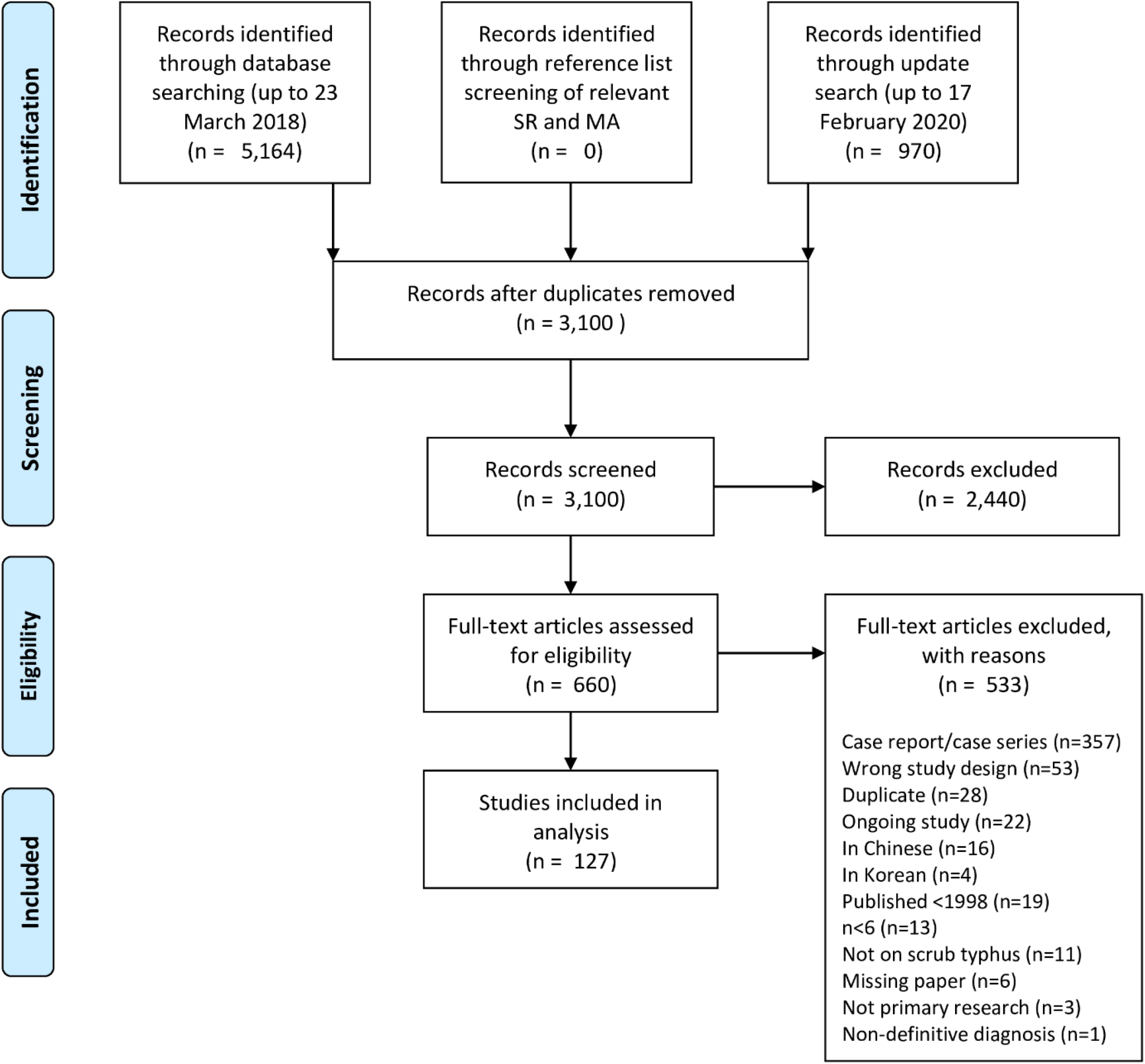
PRISMA flowchart. MA = meta-analyses, SR = systematic reviews.

### Study settings and population

The distribution of included studies is depicted in Fig 2. All studies were conducted in countries within the classic tsutsugamushi triangle. Of the 12 countries represented, India had the highest number (n= 84/127, 66%) of studies, followed by the Republic of Korea (South Korea) (n=14/127, 11%) and Thailand (n= 11/127, 9%) (Fig A, S4 Appendix). This review identified no published antibiotic treatment data for *Candidatus* Orientia chuto and *Candidatus* Orientia chiloensis.

**Fig 2.**
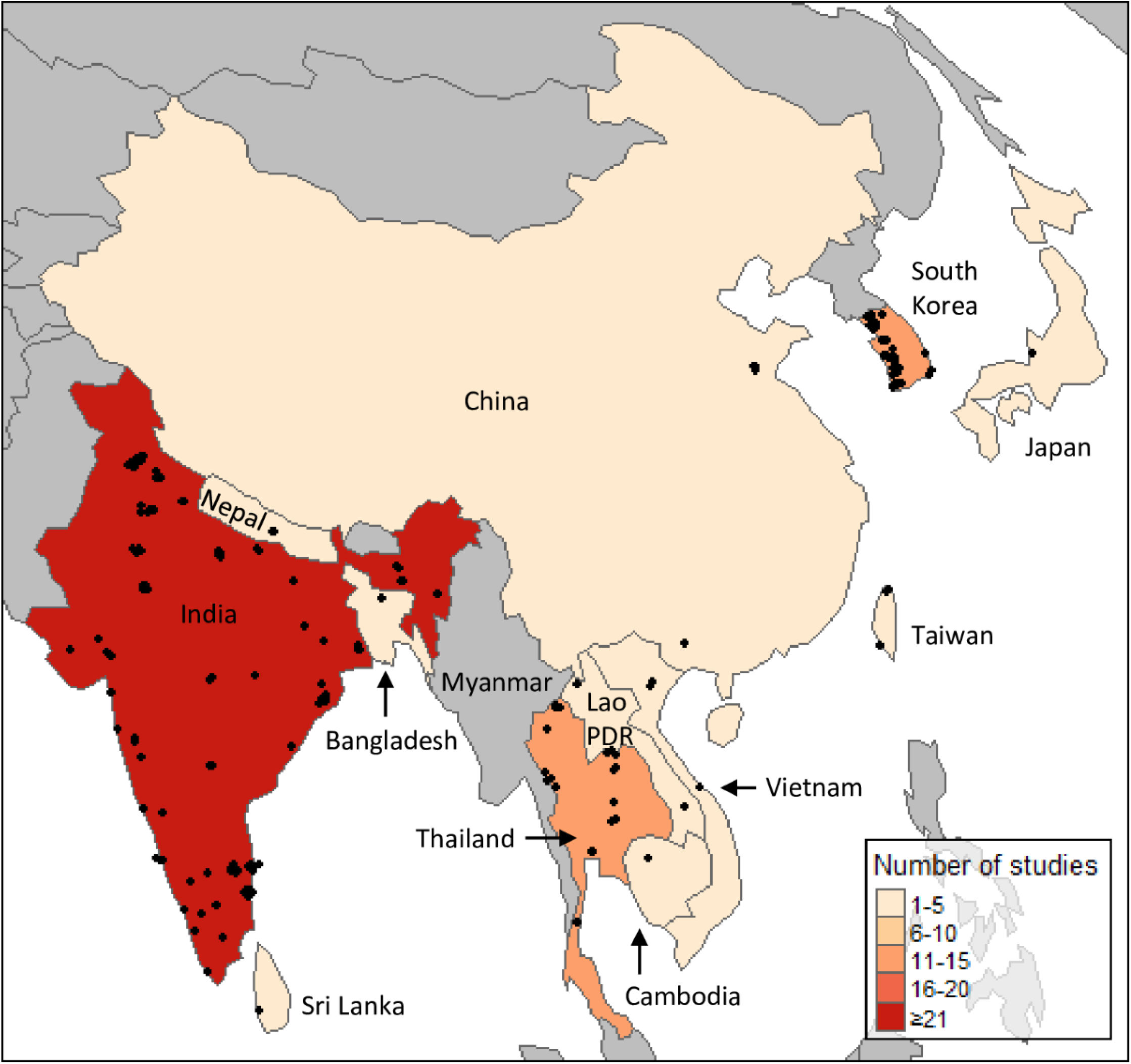
Number of eligible studies from each country. Jittered points represent each study location.

There were 141 study arms included in the 127 studies. In total, there were 12,079 patients enrolled in the studies, with most studies (n=84) and over two-thirds of participants recruited in India (n=8,377, 69%). The 14 and 11 studies conducted in South Korea and Thailand recruited 1,592 (13%) and 707 (6%) participants, respectively (Fig B, S4 Appendix). The median number of scrub typhus participants recruited in the studies was 57 (range 6-502) (Fig 3). Most of the studies (n=109/127, 86%) recruited less than 200 participants.

**Fig 3.**
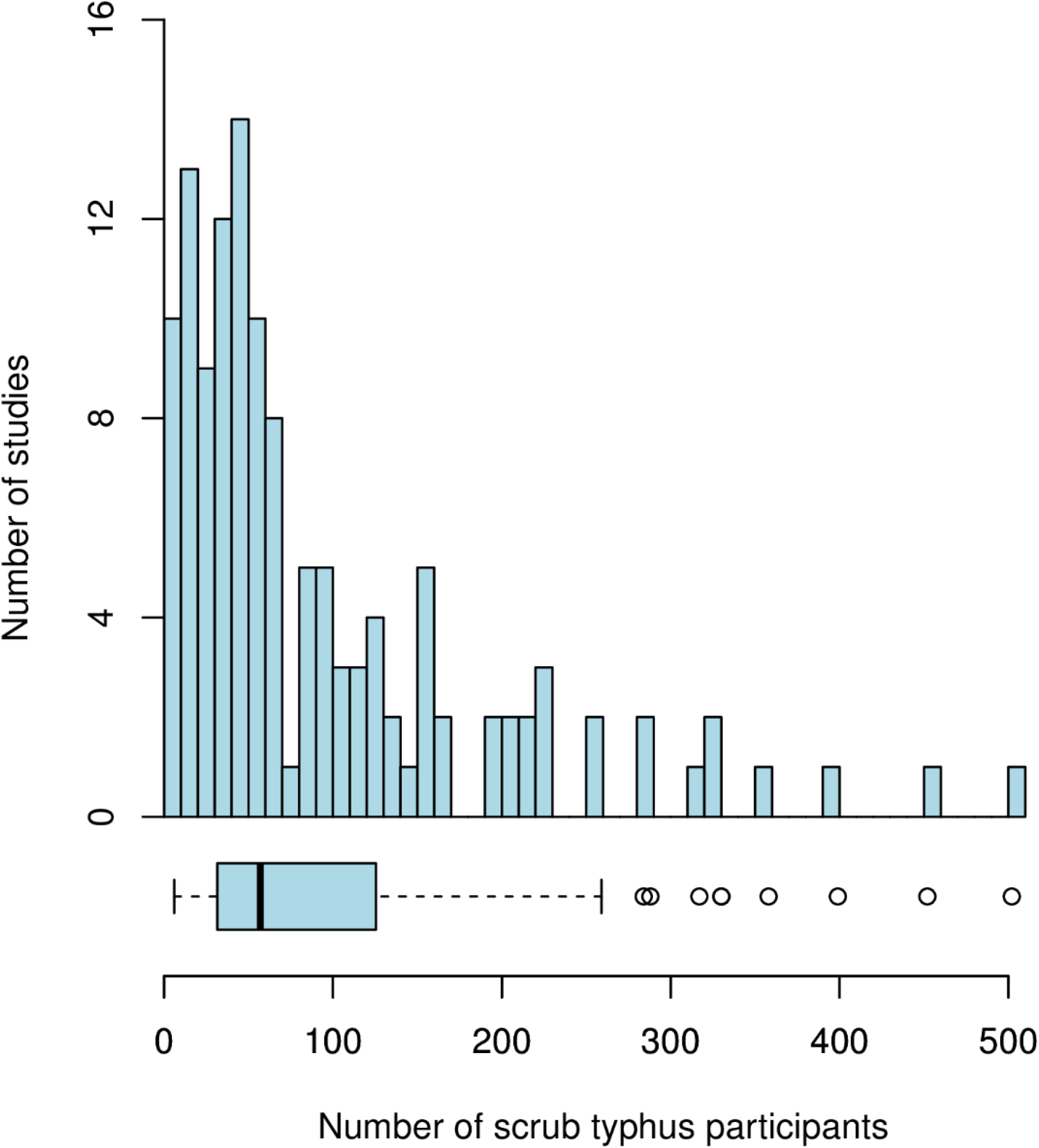
Number of scrub typhus participants enrolled per study. Bin width is 10. The box denotes the lower and upper quartiles with the median indicated by the thick vertical line. Capped bars represent the minimum and maximum values within 1.5*interquartile range of the first and third quartiles, respectively. Circles indicate values outside this interval.

The majority of the included studies were conducted in lower middle income countries as defined by the World Bank (n=98/127, 77%) (Fig C, S4 Appendix) [19]. Most studies were conducted in urban areas (n=98/127, 77%), and almost all studies recruited participants from a hospital/health-centre (n=125/127, 98%) (Fig C, S4 Appendix). One study recruited participants from a school (n=1/127, 0.8%) and one from antenatal clinics (n=1/127, 0.8%). There were no interventional studies, including pregnant women and only three interventional studies had included children. Pregnant women and children (age<18 years) were included in 14 and 75 observational studies, respectively.

### Diagnostic methods

The methods used to diagnose scrub typhus varied substantially across the included studies (Table 1). The most commonly used diagnostic method was enzyme-linked immunosorbent assay (ELISA) (n=65/127, 51%). Indirect immunofluorescence assay (IFA), which is typically considered as the gold standard, was used by 33 studies (n=33/127, 26%). Polymerase chain reaction (PCR) was used in 30 (n=30/127, 24%) studies; Weil-Felix in 25 (n=25/127, 20%); immunochromatography test in 16 (n=16/127, 13%); eschar presence in 13 (n=13/127, 10%); other clinical presentation in 6 (n=6/127, 5%); indirect immunoperoxidase test in 4 (n=4/127, 3%); cell culture isolate in 3 (n=3/127, 2%). Six (n=6/127, 5%) studies used other diagnostic methods, including hemagglutination assay (PHA), dotblot immunoassay, microimmunofluorescence, and favourable response to antibiotics. No studies used the scrub typhus infection criteria (STIC) [20]. Nine observational studies, including 186 participants, gave no details on the diagnostic methods used. For interventional studies, IFA was the most commonly used diagnostic method (n=4/7, 57%). While for observational studies, the most commonly used diagnostic method was ELISA (n=65/120, 54%). The proportion of studies using PCR and ELISA showed an increasing trend over time (Fig D, S4 Appendix).

**Table 1.**
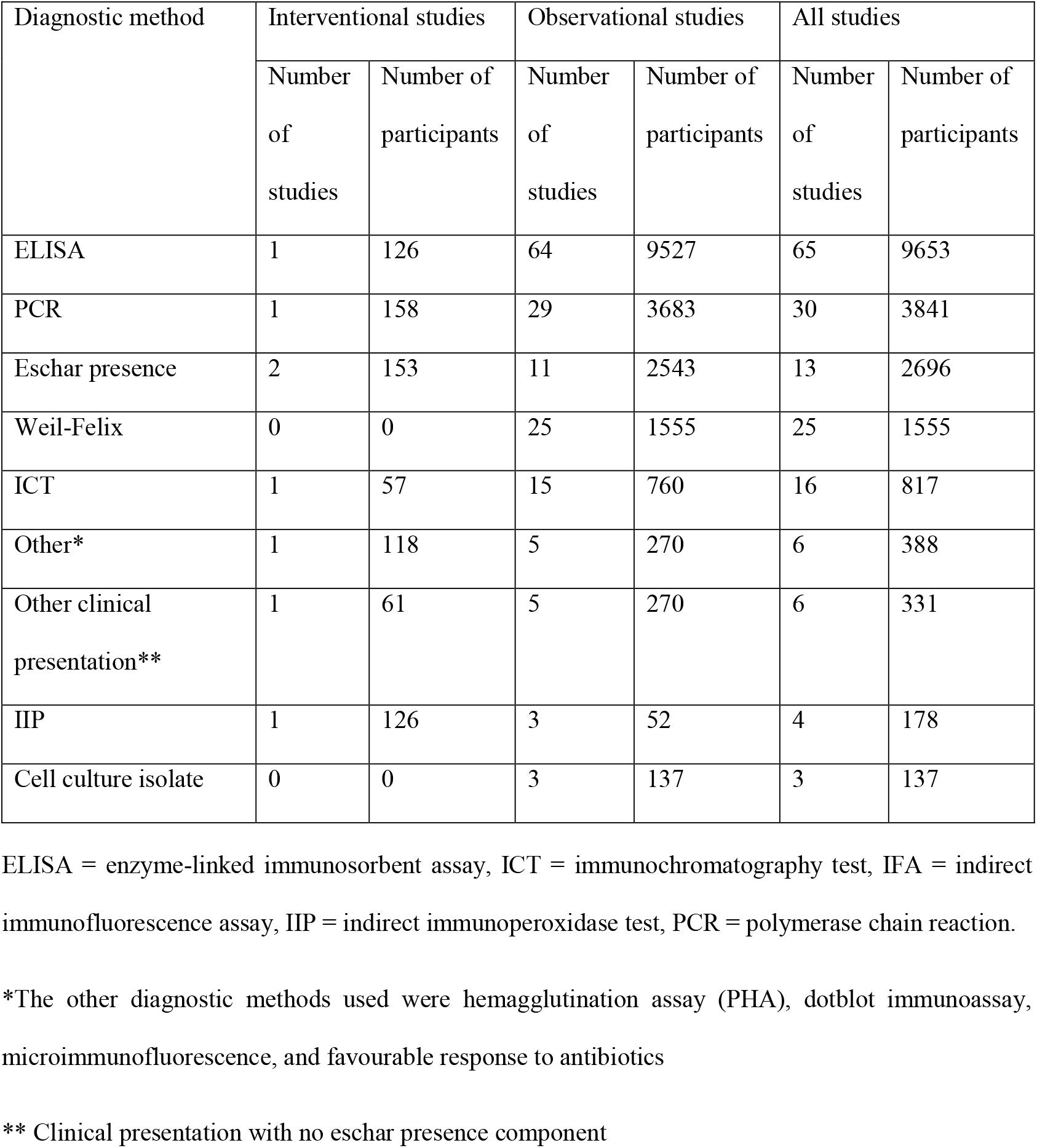
Diagnostic methods distribution across studies and participants

| Diagnostic method | Interventional studies |  | Observational studies |  | All studies |  |
| --- | --- | --- | --- | --- | --- | --- |
|  | Number of studies | Number of participants | Number of studies | Number of participants | Number of studies | Number of participants |
| ELISA | 1 | 126 | 64 | 9527 | 65 | 9653 |
| PCR | 1 | 158 | 29 | 3683 | 30 | 3841 |
| Eschar presence | 2 | 153 | 11 | 2543 | 13 | 2696 |
| Weil-Felix | 0 | 0 | 25 | 1555 | 25 | 1555 |
| ICT | 1 | 57 | 15 | 760 | 16 | 817 |
| Other* | 1 | 118 | 5 | 270 | 6 | 388 |
| Other clinical presentation** | 1 | 61 | 5 | 270 | 6 | 331 |
| IIP | 1 | 126 | 3 | 52 | 4 | 178 |
| Cell culture isolate | 0 | 0 | 3 | 137 | 3 | 137 |

ELISA = enzyme-linked immunosorbent assay, ICT = immunochromatography test, IFA = indirect immunofluorescence assay, IIP = indirect immunoperoxidase test, PCR = polymerase chain reaction.
\*The other diagnostic methods used were hemagglutination assay (PHA), dotblot immunoassay, microimmunofluorescence, and favourable response to antibiotics
\*\* Clinical presentation with no eschar presence component

### Treatment

There was substantial heterogeneity in treatment, with 14 types of antibiotics administered in the included studies. Doxycycline was the most commonly administered antibiotic (94/141 study arms with 8,062 participants), followed by azithromycin and chloramphenicol, which were used in 55 (5,855 participants) and 20 (1,801 participants) study arms, respectively (Fig 4). All other antibiotics were administered to less than ten study arms and less than 1,000 participants.

**Fig 4.**
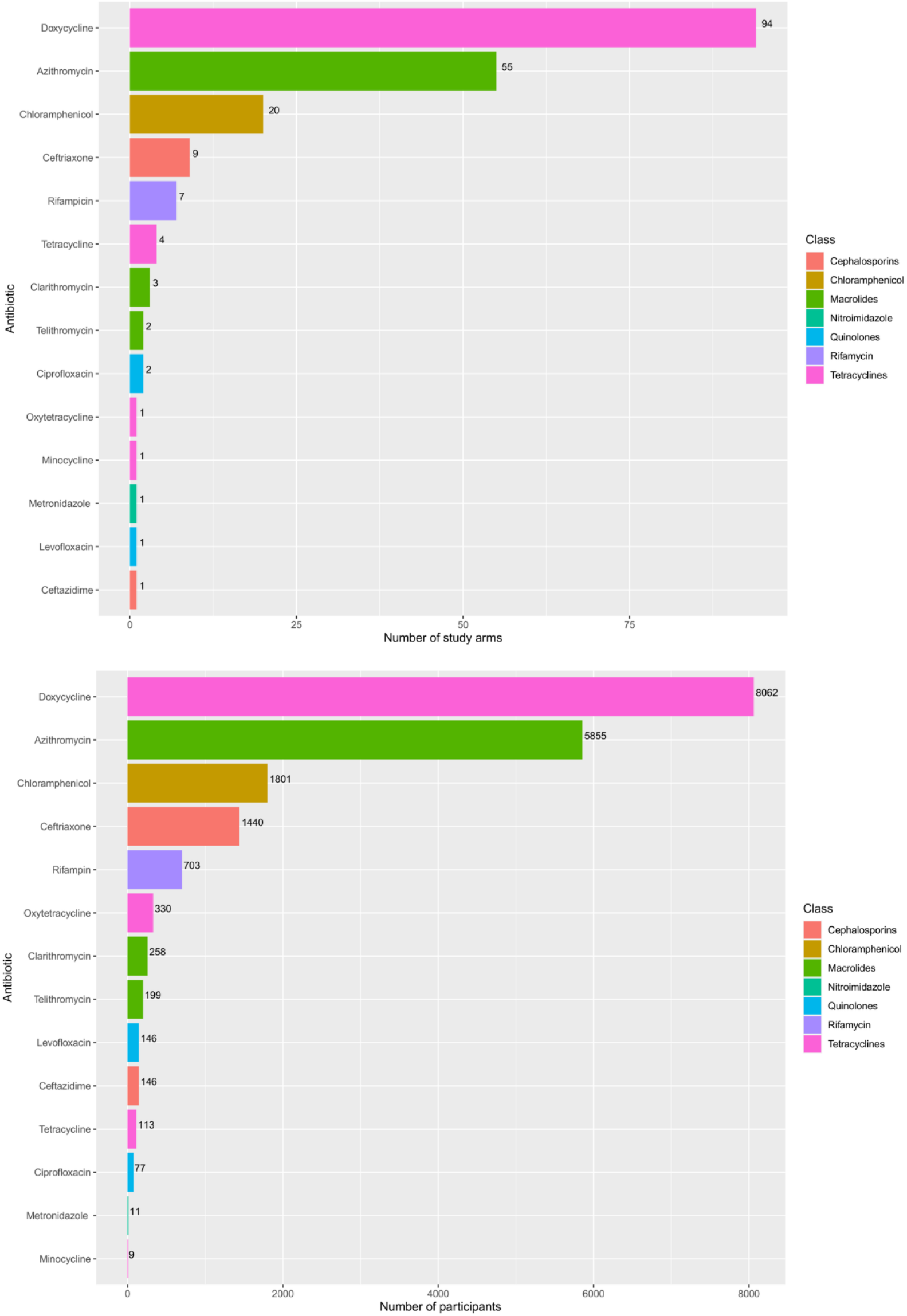
Number of study arms and participants within studies administering each antibiotic.

The dosage regimen administered in the studies varied substantially. We identified 52 described combinations of drug, dose, and frequency. When the duration of antibiotic treatment was also considered, there were 66 unique combinations of drug, dose, frequency and duration. There were ten dosage regimens reported for doxycycline: one with loading dose and nine with no loading dose. Two study arms administered 200 mg doxycycline followed by a 7-day course of 100 mg twice daily or every 12 hours. Among the reported dosage regimens with no loading dose, the most often used doxycycline regimen was 100 mg twice daily (n=23/35, 64%) (Fig 5). There were six dosage regimens reported for azithromycin: two with loading dose and four with no loading dose. One study arm used 20 mg/kg/dose loading dose, followed by 10 mg/kg/dose for two days. One study arm administered 1 g followed by 500 mg once daily for two days. Within the dosage regimens without loading dose, the most commonly administered azithromycin regimen was 500 mg once daily (n=7/13, 54%) (Fig 5). For chloramphenicol, there were three identified dosage regimens (administered in one study arm each): 500 mg four times daily, 500-1000 mg every six hours, and 25-50 mg/kg/day four times daily.

**Fig 5.**
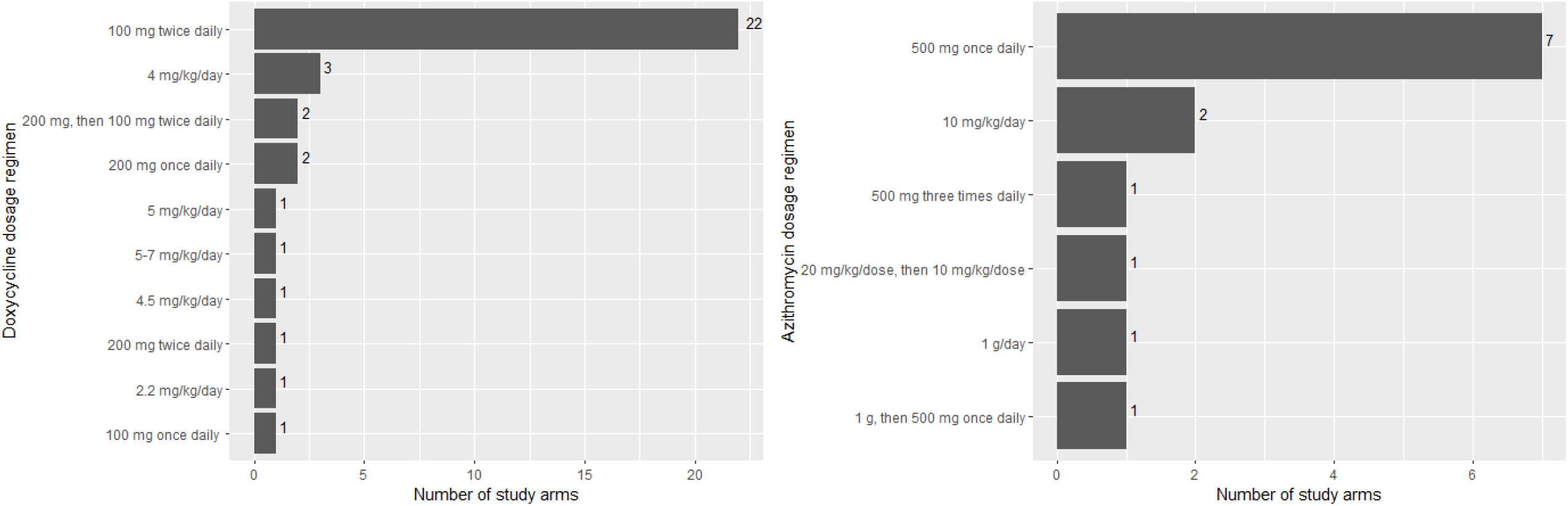
Number of study arms administering each doxycycline and azithromycin dosage regimen

### Patient outcomes

Mortality, complications, adverse events and clinical responses were assessed in the majority of studies with 106 (10,794 participants), 99 (10,103 participants), 93 (9,688 participants), and 87 studies (7,895 participants) reporting these outcomes, respectively (Fig 6, Table in S4 Appendix). They were also the most commonly assessed outcomes among the observational studies with 100 (10,211 participants), 95 (9,770 participants), 87 (9,105 participants) and 80 (7,251 participants) studies, respectively. Conversely, clinical response, fever clearance, and fever clearance time were the most commonly assessed outcomes among interventional studies (7 studies each, 644 participants).

**Fig 6.**
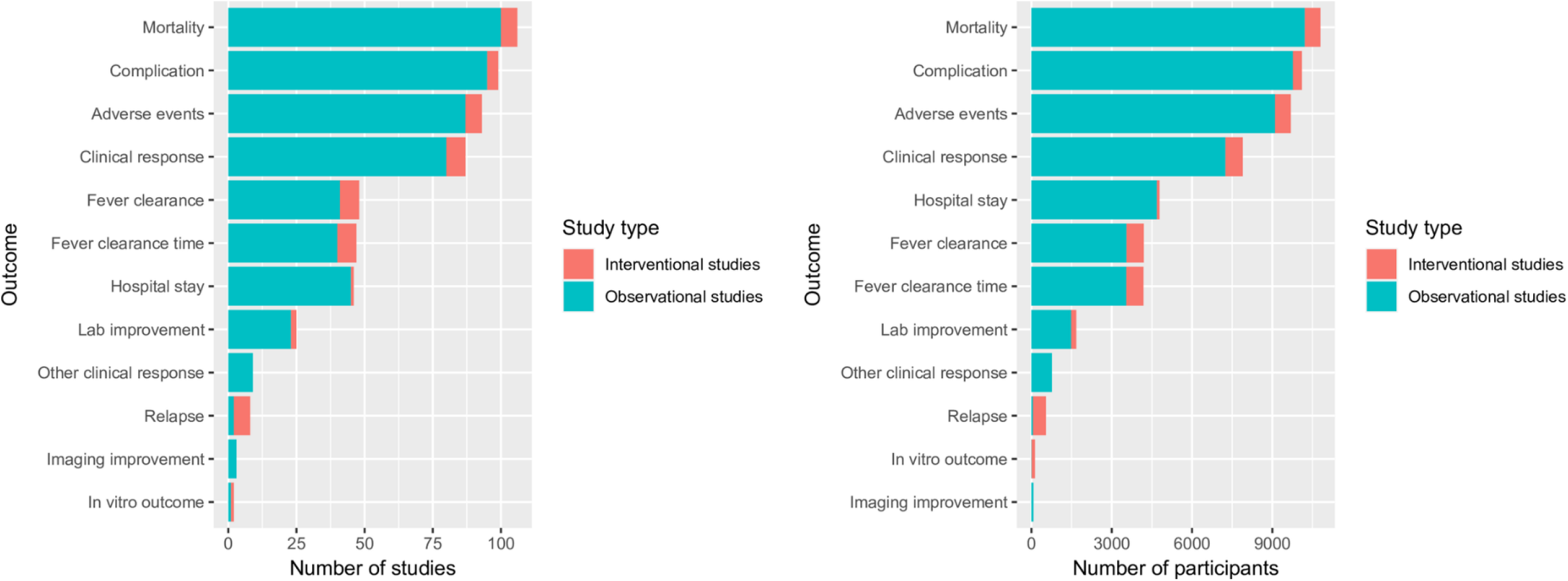
Number of studies and participants within studies assessing each outcomes

The interventional studies used varying definition of fever clearance time and relapse. Thirty-nine out of 47 studies recording fever clearance time (FCT) as an outcome did not specify the definition of FCT. There was substantial variability in the definition of FCT in the other eight studies concerning temperature cut-off, treatment, antibiotic, antipyretic, time (within 24-hour and 48-hour) and two times temperature measurement to confirm fever clearance (Table B, S4). Relapse was defined in two out of seven studies noting this outcome; both studies defined relapse as the reappearance of fever and other scrub typhus signs and symptoms within one month (30 days) in the absence of other causes.

## Discussion

After screening 3,100 records, we identified and reviewed 127 scrub typhus treatment studies comprised of 141 study arms (referring both to a study arm from a clinical trial or a patient cohort from an observational study where treatment is administered) and 12,079 participants. All of these studies were conducted within the classical tsutsugamushi triangle. However, in concordance with the findings of another review conducted by the Infectious Diseases Data Observatory (IDDO) [21], it is worth noting that even within the tsutsugamushi triangle, information is lacking – many countries did not have any data points, e.g. Myanmar and Pakistan. Substantial heterogeneity was observed in diagnosis methods, antibiotic treatment regimens, and observed outcomes.

Despite being one of the most commonly reported pathogens in South and South-East Asia [21], the review illustrates the limited number of interventional studies and very sparse data collected on specific groups such as children and pregnant women, reflecting that scrub typhus is indeed a neglected disease in terms of research to inform policy. Other studies have also shown that scrub typhus trials have a high risk of bias as assessed by the Cochrane risk of bias tool [7, 11]. These findings illustrate specific knowledge gaps which prospective studies could cover.

However, if assembled, the existing data could answer important research questions through IPD meta-analyses. Mortality, adverse events, complications, and clinical response were captured by the majority of studies, raising the possibility that these studies could be combined and analysed. The creation of an IPD platform would allow some level of standardisation between studies which would be critical to facilitate pooling of available data and account for the heterogeneity of these studies [22, 23].

To our knowledge, this is the first systematic review exploring the scrub typhus treatment study landscape. We conducted an extensive search in electronic databases and clinical trial registries. We also included observational studies to ensure a more comprehensive identification of IPD. We flagged ongoing studies to allow evaluation of what kind of data would be available in the future. Although no language restrictions were applied during the search, we could not extract data from articles in Korean and Chinese due to limited translation resources. This review only included peer-reviewed published studies. We also specified the search to start in 1998 due to the small probability of obtaining individual patient data from earlier studies [15]. This process might introduce selection bias, and there may be more IPD available in ongoing investigations and grey literature such as government surveillance data that were not identified in this search. Due to resources restrictions, only one reviewer screened the title and abstract. This might introduce classification error in the selection stage. However, we were able to deploy two reviewers in the full-text screening and data extraction stage.

This review highlights the heterogeneity in the methods and reporting of scrub typhus treatment studies. There are various treatment regimens administered and outcomes observed. Due to differences in definitions used, it would be challenging to conduct a useful aggregated data meta-analysis without strong assumptions and access to additional data and information. In this situation, curation and standardisation of IPD would help in pooling available data [22, 23].

In the studies we examined there were 7,835 participants treated with doxycycline, 5,970 with azithromycin, and 2,056 with chloramphenicol – which should be sufficient to estimate their efficacy and potential patient-level factors affecting the outcomes.

To optimise the value and utility of data gathered through clinical research, it would be valuable to define a set of data and design standards for RCTs and observational studies to facilitate comparisons across studies. Standards of data collected should include definition of key variables, study design, diagnostic methods, and endpoints. More importantly, these standards need to be implemented consistently. Furthermore, it is essential to adhere to reporting guidelines such as the STROBE (STrengthening the Reporting of OBservational studies in Epidemiology) and CONSORT (Consolidated Standards of Reporting Trials) checklists during design and publication [24, 25].

The Clinical Data Interchange Standards Consortium (CDISC) develops data standards to ensure clarity and maximise the interoperability and reusability of data, consequently improving data value and impact. It is required by many regulatory authorities and leading research organisations [26]. This would provide a framework for collecting, organising, analysing, and sharing data. This framework, complemented by consensus on outcomes, can ensure comparability and interoperability of data. The COMET (Core Outcome Measures in Effectiveness Trials) Initiative has compiled resources to develop and apply core outcome sets (COS) for clinical trials [27]. COS is ‘an agreed standardised set of outcomes that should be measured and reported, as a minimum, in all clinical trials in specific areas of health or health care’ [27]. Such an approach is needed in scrub typhus clinical trials and observational studies to facilitate meta-analysis and comparisons between studies. Concerted efforts from stakeholders will be needed to manifest this.

In the future, further quality assessments of clinical trials may be useful to identify the source of heterogeneity and identify any barriers to develop more standardised methods and reporting. Formulation of standardised definition, procedures to assess treatment efficacy, and reporting through a consensus-based approach within the research community is an important next step. For data sharing to become normalised, mechanisms need to be put in place to ensure a mutually beneficial relationship between people who re-use the data and those who collect them [23]. Engagement and cooperation with the scrub typhus research community and investment in data infrastructure to implement standards and facilitate data sharing and meta-analysis are of utmost importance to establish an IPD platform.

One way to increase awareness of scrub typhus is its recognition as an NTD by the WHO. Scrub typhus fulfils three out of the four WHO suggested criteria for NTD [28]: (1) it causes ‘important morbidity and mortality’ particularly in rural population with a higher rate of poverty and less access to health care facilities, (2) affecting people in tropical and subtropical areas, and (3) ‘neglected by research’. The inclusion of scrub typhus would prompt support for advocacy and development of diagnostics tools, treatment, and control strategies [28]. When ‘effective and feasible’ control strategies are available, scrub typhus can fulfil the last criteria – a disease that can be immediately controlled with public health interventions [28, 29].

There is still considerable uncertainty about the optimal treatment for scrub typhus, reflected by the highly heterogeneous set of treatment regimens used in the studies reviewed over the last 22 years. Despite the low number of scrub typhus treatment trials published, there is likely a reasonable amount of data available with key information to address this uncertainty. This review further supports the case for the development of a scrub typhus IPD platform. This will facilitate pooled analysis of treatment data that will in turn, inform clinical practice and disease control measures. Stakeholder engagement and cooperation will be crucial to the establishment of a scrub typhus IPD platform.

## Supporting information

S1 Checklist. PRISMA checklist

S2 Appendix. Search strategy

S3 Appendix. List of included and excluded studies

S4 Appendix. Additional results

S5 Appendix. Variable dictionary

S6 Appendix. Data dictionary

## Data Availability

All relevant data are within the manuscript and its Supporting Information files.

## List of supporting information

S1 Checklist. PRISMA checklist

S2 Appendix. Search strategy

S3 Appendix. List of included and excluded studies

S4 Appendix. Additional results

S5 Appendix. Variable dictionary

S6 Appendix. Data dictionary

## Acknowledgement

We would like to thank Ms Elinor Harriss, Outreach Librarian, Bodleian Health Care Libraries, University of Oxford and Ms Kalynn Kennon, Head of Data Engineering, Infectious Diseases Data Observatory for their support throughout this work.

## Data availability statement

All relevant data are within the manuscript and its Supporting Information files.

## Authors Contributions

Conceptualisation: PN, ND, PG

Data Curation: KS, SS, RN, BM

Formal Analysis: KS, BM, AM

Funding Acquisition: PN, ND, PG

Investigation: KS, SS, RN, BM, AM

Methodology: KS, BM, ND, PG

Project Administration: KS

Resources: N/A

Software: KS, AM

Supervision: ND, PG

Validation: KS, SS, RN, BM

Visualisation: KS, BM, AM

Writing – Original Draft Preparation: KS, BM, AM

Writing – Review & Editing: KS, BM, AM, SS, RN, PN, ND, PG

## Conflict of Interest

All authors declare no competing interests.

## Funding

The review was funded by a biomedical resource grant from Wellcome (https://wellcome.org/) to the Infectious Diseases Data Observatory (Recipient: PJG; ref: 208378/Z/17/Z). The funders had no role in study design, data collection and analysis, decision to publish, or preparation of the manuscript.

## Notes

### Competing Interest Statement

The authors have declared no competing interest.

### Summary of Updates

Data availability statement updated since the webpage link is not currently live.

