## Supplementary material for "Systematic review of the scrub typhus treatment landscape: Assessing the feasibility of an individual participant-level data (IPD) platform": S2 Appendix. Search strategy

### Search strategies by electronic bibliographic database

#### Ovid (Embase)

Database: Embase

Search Strategy:

--------------------------------------------------------------------------------

1 scrub typhus/

2 Orientia tsutsugamushi/

3 "scrub typhus".ti,ab.

4 tsutsugamushi.ti,ab.

5 scrubtyphus.mp. [mp=title, abstract, heading word, drug trade name, original title, device manufacturer, drug manufacturer, device trade name, keyword, floating subheading word]

6 "mite typhus".mp. [mp=title, abstract, heading word, drug trade name, original title, device manufacturer, drug manufacturer, device trade name, keyword, floating subheading word]

7 "japanese river fever".mp. [mp=title, abstract, heading word, drug trade name, original title, device manufacturer, drug manufacturer, device trade name, keyword, floating subheading word]

8 "kedani fever".mp. [mp=title, abstract, heading word, drug trade name, original title, device manufacturer, drug manufacturer, device trade name, keyword, floating subheading word]

9 "chigger borne rickettsia".mp. [mp=title, abstract, heading word, drug trade name, original title, device manufacturer, drug manufacturer, device trade name, keyword, floating subheading word]

10 "chigger borne rickettsiosis".mp. [mp=title, abstract, heading word, drug trade name, original title, device manufacturer, drug manufacturer, device trade name, keyword, floating subheading word]

11 "chigger borne typhus".mp. [mp=title, abstract, heading word, drug trade name, original title, device manufacturer, drug manufacturer, device trade name, keyword, floating subheading word]

12 mijtekoorts.mp. [mp=title, abstract, heading word, drug trade name, original title, device manufacturer, drug manufacturer, device trade name, keyword, floating subheading word]

13 "mite borne rickettsia".mp. [mp=title, abstract, heading word, drug trade name, original title, device manufacturer, drug manufacturer, device trade name, keyword, floating subheading word]

14 "mite borne rickettsiosis".mp. [mp=title, abstract, heading word, drug trade name, original title, device manufacturer, drug manufacturer, device trade name, keyword, floating subheading word]

15 "mite borne typhus".mp. [mp=title, abstract, heading word, drug trade name, original title, device manufacturer, drug manufacturer, device trade name, keyword, floating subheading word]

16 1 or 2 or 3 or 4 or 5 or 6 or 7 or 8 or 9 or 10 or 11 or 12 or 13 or 14 or 15

17 limit 16 to humans

18 limit 17 to yr="1998 -Current"

19 limit 18 to editorial

20 limit 18 to letter

21 limit 18 to "review"

22 limit 18 to "systematic review"

23 limit 18 to meta analysis

24 19 or 20 or 21 or 22 or 23

25 18 not 24

#### Ovid (Medline)

Database: Ovid MEDLINE(R) Epub Ahead of Print, In-Process & Other Non-Indexed Citations, Ovid MEDLINE(R) Daily, Ovid MEDLINE and Versions(R)

Search Strategy:

--------------------------------------------------------------------------------

1 scrub typhus/

2 Orientia tsutsugamushi/

3 "scrub typhus".ti,ab.

4 tsutsugamushi.ti,ab.

5 scrubtyphus.mp. [mp=title, abstract, original title, name of substance word, subject heading word, keyword heading word, protocol supplementary concept word, rare disease supplementary concept word, unique identifier, synonyms]

6 "mite typhus".mp. [mp=title, abstract, original title, name of substance word, subject heading word, keyword heading word, protocol supplementary concept word, rare disease supplementary concept word, unique identifier, synonyms]

7 "japanese river fever".mp. [mp=title, abstract, original title, name of substance word, subject heading word, keyword heading word, protocol supplementary concept word, rare disease supplementary concept word, unique identifier, synonyms]

8 "kedani fever".mp. [mp=title, abstract, original title, name of substance word, subject heading word, keyword heading word, protocol supplementary concept word, rare disease supplementary concept word, unique identifier, synonyms]

9 "chigger borne rickettsia".mp. [mp=title, abstract, original title, name of substance word, subject heading word, keyword heading word, protocol supplementary concept word, rare disease supplementary concept word, unique identifier, synonyms]

10 "chigger borne rickettsiosis".mp. [mp=title, abstract, original title, name of substance word, subject heading word, keyword heading word, protocol supplementary concept word, rare disease supplementary concept word, unique identifier, synonyms]

11 "chigger borne typhus".mp. [mp=title, abstract, original title, name of substance word, subject heading word, keyword heading word, protocol supplementary concept word, rare disease supplementary concept word, unique identifier, synonyms]

12 mijtekoorts.mp. [mp=title, abstract, original title, name of substance word, subject heading word, keyword heading word, protocol supplementary concept word, rare disease supplementary concept word, unique identifier, synonyms]

13 "mite borne rickettsia".mp. [mp=title, abstract, original title, name of substance word, subject heading word, keyword heading word, protocol supplementary concept word, rare disease supplementary concept word, unique identifier, synonyms]

14 "mite borne rickettsiosis".mp. [mp=title, abstract, original title, name of substance word, subject heading word, keyword heading word, protocol supplementary concept word, rare disease supplementary concept word, unique identifier, synonyms]

15 "mite borne typhus".mp. [mp=title, abstract, original title, name of substance word, subject heading word, keyword heading word, protocol supplementary concept word, rare disease supplementary concept word, unique identifier, synonyms]

16 1 or 2 or 3 or 4 or 5 or 6 or 7 or 8 or 9 or 10 or 11 or 12 or 13 or 14 or 15

17 limit 16 to human

18 limit 17 to yr="1998 -Current"

19 limit 18 to systematic reviews

20 limit 18 to meta analysis

21 limit 18 to editorial

22 limit 18 to letter

23 limit 18 to "review"

24 19 or 20 or 21 or 22 or 23

25 18 not 24

#### Ovid (Global Health)

Database: Global Health

Search Strategy:

--------------------------------------------------------------------------------

scrub typhus/

Orientia tsutsugamushi/

"scrub typhus".ti,ab.

tsutsugamushi.ti,ab.

scrubtyphus.mp. [mp=abstract, title, original title, broad terms, heading words, identifiers, cabicodes]

"mite typhus".mp. [mp=abstract, title, original title, broad terms, heading words, identifiers, cabicodes]

"japanese river fever".mp. [mp=abstract, title, original title, broad terms, heading words, identifiers, cabicodes]

"kedani fever".mp. [mp=abstract, title, original title, broad terms, heading words, identifiers, cabicodes]

"chigger borne rickettsia".mp. [mp=abstract, title, original title, broad terms, heading words, identifiers, cabicodes]

"chigger borne rickettsiosis".mp. [mp=abstract, title, original title, broad terms, heading words, identifiers, cabicodes]

"chigger borne typhus".mp. [mp=abstract, title, original title, broad terms, heading words, identifiers, cabicodes]

mijtekoorts.mp. [mp=abstract, title, original title, broad terms, heading words, identifiers, cabicodes]

"mite borne rickettsia".mp. [mp=abstract, title, original title, broad terms, heading words, identifiers, cabicodes]

"mite borne rickettsiosis".mp. [mp=abstract, title, original title, broad terms, heading words, identifiers, cabicodes]

"mite borne typhus".mp. [mp=abstract, title, original title, broad terms, heading words, identifiers, cabicodes]

1 or 2 or 3 or 4 or 5 or 6 or 7 or 8 or 9 or 10 or 11 or 12 or 13 or 14 or 15

"systematic review".mp. and 16 [mp=abstract, title, original title, broad terms, heading words, identifiers, cabicodes]

("meta analysis" or "meta-analysis").mp. and 16 [mp=abstract, title, original title, broad terms, heading words, identifiers, cabicodes]

limit 16 to yr="1998 -Current"

limit 17 to yr="1998 -Current"

limit 19 to yr="1998 -Current"

#### Cochrane library

"scrub typhus" OR tsutsugamushi OR scrubtyphus OR "mite typhus" OR "japanese river fever" OR "kedani fever" OR "chigger borne rickettsia" OR "chigger borne rickettsiosis" OR "chigger borne typhus" OR mijtekoorts OR "mite borne rickettsia" OR "mite borne rickettsiosis" OR "mite borne typhus"

Publication Year from 1998 to 2018

#### Scopus

( TITLE-ABS-KEY ( "scrub typhus" ) OR TITLE-ABS-KEY ( tsutsugamushi ) OR ALL ( scrubtyphus ) OR ALL ( "mite typhus" ) OR ALL ( "japanese river fever" ) OR ALL ( "kedani fever" ) OR ALL ( "chigger borne rickettsia" ) OR ALL ( "chigger borne rickettsiosis" ) OR ALL ( "chigger borne typhus" ) OR ALL ( mijtekoorts ) OR ALL ( "mite borne rickettsia" ) OR ALL ( "mite borne rickettsiosis" ) OR ALL ( "mite borne typhus" ) ) AND PUBYEAR > 1997 AND ( LIMIT-TO ( DOCTYPE , "ar" ) OR LIMIT-TO ( DOCTYPE , "cp" ) OR LIMIT-TO ( DOCTYPE , "ip" ) )

#### Global Index Medicus

tw:("scrub typhus" OR tsutsugamushi OR scrubtyphus OR "mite typhus" OR "japanese river fever" OR "kedani fever" OR "chigger borne rickettsia" OR "chigger borne rickettsiosis" OR "chigger borne typhus" OR mijtekoorts OR "mite borne rickettsia" OR "mite borne rickettsiosis" OR "mite borne typhus") AND (instance:"ghl") AND ( db:("WPRIM") AND year_cluster:("1998"or "1999"or "2000"or "2001"or "2002"or "2003"or "2004"or "2005"or "2006"or "2007"or "2008"or "2009"or "2010"or "2011"or "2012"or "2013"or "2014"or "2015"or "2016"or "2017"or "2018"))

tw:("scrub typhus" OR tsutsugamushi OR scrubtyphus OR "mite typhus" OR "japanese river fever" OR "kedani fever" OR "chigger borne rickettsia" OR "chigger borne rickettsiosis" OR "chigger borne typhus" OR mijtekoorts OR "mite borne rickettsia" OR "mite borne rickettsiosis" OR "mite borne typhus") AND (instance:"ghl") AND ( db:("IMSEAR") AND year_cluster:("1998"or "1999"or "2000"or "2001"or "2002"or "2003"or "2004"or "2005"or "2006"or "2007"or "2008"or "2009"or "2010"or "2011"or "2012"or "2013"or "2014"or "2015"or "2016"or "2017"or "2018"))

tw:("scrub typhus" OR tsutsugamushi OR scrubtyphus OR "mite typhus" OR "japanese river fever" OR "kedani fever" OR "chigger borne rickettsia" OR "chigger borne rickettsiosis" OR "chigger borne typhus" OR mijtekoorts OR "mite borne rickettsia" OR "mite borne rickettsiosis" OR "mite borne typhus") AND (instance:"ghl") AND ( db:("LILACS") AND year_cluster:("1998"or "1999"or "2000"or "2001"or "2002"or "2003"or "2004"or "2005"or "2006"or "2007"or "2008"or "2009"or "2010"or "2011"or "2012"or "2013"or "2014"or "2015"or "2016"or "2017"or "2018"))

tw:("scrub typhus" OR tsutsugamushi OR scrubtyphus OR "mite typhus" OR "japanese river fever" OR "kedani fever" OR "chigger borne rickettsia" OR "chigger borne rickettsiosis" OR "chigger borne typhus" OR mijtekoorts OR "mite borne rickettsia" OR "mite borne rickettsiosis" OR "mite borne typhus") AND (instance:"ghl") AND ( db:("IMEMR") AND year_cluster:("1998"or "1999"or "2000"or "2001"or "2002"or "2003"or "2004"or "2005"or "2006"or "2007"or "2008"or "2009"or "2010"or "2011"or "2012"or "2013"or "2014"or "2015"or "2016"or "2017"or "2018"))

#### ClinicalTrials.gov

"scrub typhus"

#### WHO ICTRP

"scrub typhus"
