## Supplementary material for "Systematic review of the scrub typhus treatment landscape: Assessing the feasibility of an individual participant-level data (IPD) platform": S4 Appendix. Additional results

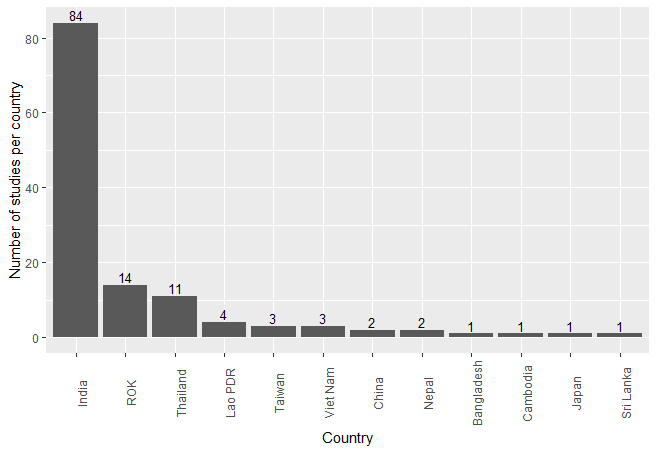

Figure A. Number of studies per country. Lao PDR = Lao People Democratic Republic, ROK = Republic of Korea (South Korea).

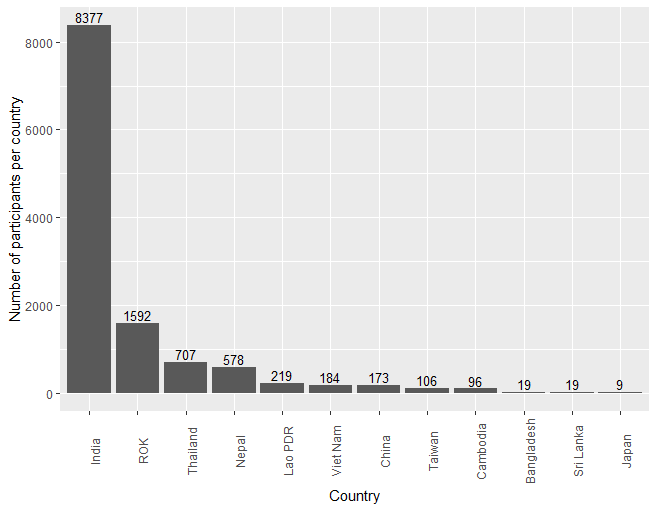

Figure B. Number of participants per country. Lao PDR = Lao People Democratic Republic, ROK = Republic of Korea (South Korea).

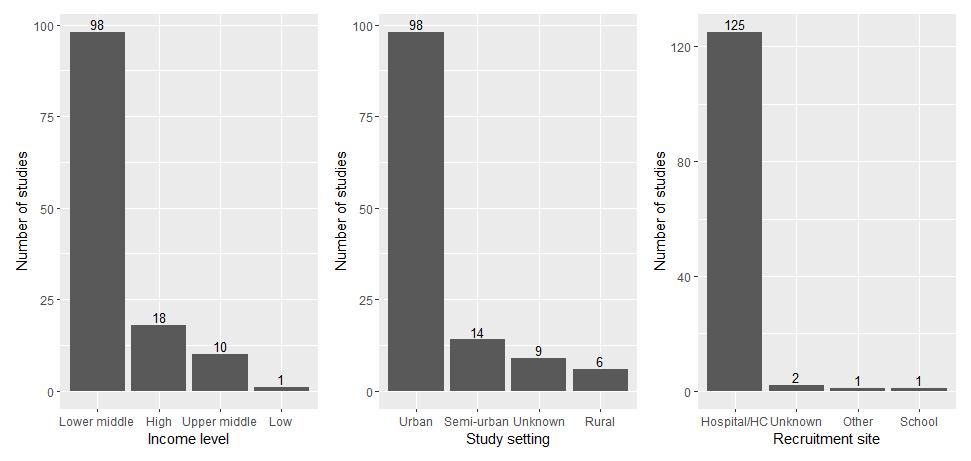

Figure C. Number of studies for each income level, study setting, and recruitment site

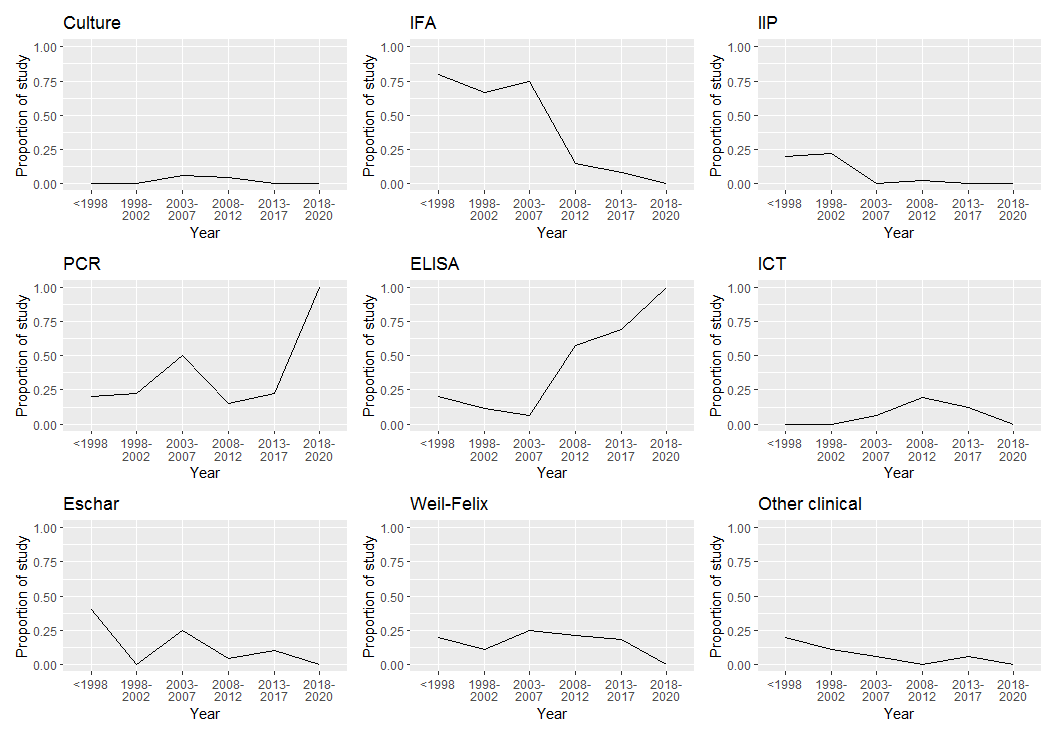

Figure D. The proportion of studies using each diagnostic methods divided by the number of studies in each year group

Table A. Number of studies and participants within studies reporting each outcome

| Outcome | Interventional studies | | Observational studies | | All studies | |
| --- | --- | --- | --- | --- | --- | --- |
|  | Number of studies | Number of participants | Number of studies | Number of participants | Number of studies | Number of participants |
| Mortality | 6 | 583 | 100 | 10211 | 106 | 10794 |
| Complication | 4 | 333 | 95 | 9770 | 99 | 10103 |
| Adverse events | 6 | 583 | 87 | 9105 | 93 | 9688 |
| Clinical response | 7 | 644 | 80 | 7251 | 87 | 7895 |
| Hospital stay | 1 | 93 | 45 | 4689 | 46 | 4782 |
| Fever clearance | 7 | 644 | 41 | 3554 | 48 | 4198 |
| Fever clearance time | 7 | 644 | 40 | 3543 | 47 | 4187 |
| Improvement of lab parameters | 2 | 185 | 23 | 1488 | 25 | 1673 |
| Clinical response besides fever clearance | 0 | 0 | 9 | 767 | 9 | 767 |
| Relapse | 6 | 486 | 2 | 58 | 8 | 544 |
| In vitro outcome | 1 | 126 | 1 | 10 | 2 | 136 |
| Imaging improvement | 0 | 0 | 3 | 77 | 3 | 77 |
| Pharmacokinetics | 0 | 0 | 0 | 0 | 0 | 0 |

Table B. The definitions of fever clearance time within studies

| Stated definition of fever clearance time* | Temperature cut-off | Treatment administration | Antibiotic administration | Antipyretic administration | 24-hour window | 48-hour window | Twice temperature measurement | Number of studies |
| --- | --- | --- | --- | --- | --- | --- | --- | --- |
| Not specified clearly |  |  |  |  |  |  |  | 39 |
| “Time to defervescence was defined as the interval between the time at which the first dose of antibiotic was administered and the time at which the body temperature was less than 37.3°C and persisted for more than 48 hours. Temperature was checked every 4 hours.” | 37.3 | 0 | 0 | 0 | 0 | 1 | 0 | 1 |
| “The primary endpoint was the fever clearance time; this was defined as the interval between the time at which the first dose of antibiotic was administered and the time at which the oral temperature first fell below 37.3°C and then remained below this threshold without antipyretics for a minimum of 48 h.” | 37.3 | 0 | 0 | 1 | 0 | 1 | 0 | 1 |
| “The time to fever clearance was defined as the period from the initial drug administration until the body temperature decreased to 37.3°C or lower and remained below this temperature for at least 48 hours without the use of an antipyretic.” | 37.3 | 1 | 0 | 1 | 0 | 1 | 0 | 2 |
| “Fever clearance time was defined as the time, from onset of treatment, to the first time the aural temperature dropped below 37.5°C and stayed at or below 37.5°C for 24 hours.” | 37.5 | 1 | 0 | 0 | 1 | 0 | 0 | 1 |
| “Time to defervescence, which was defined as the interval between the time at which the first dose of the study drug was administered and the time at which the oral temperature first returned to ≤37.5°C and was maintained for two consecutive measurements without antipyretics.” | 37.5 | 1 | 0 | 0 | 0 | 0 | 1 | 1 |
| “The fever clearance time was the interval between the first dose of antibiotic and the time when the patient's oral temperature first fell below 37·3º C, and remained below this threshold without antipyretics for a minimum of 48 h.” | 37.3 | 0 | 1 | 1 | 0 | 1 | 0 | 1 |
| “The fever clearance time was the number of hours between the first dose of medication and the first time when the morning oral temperature was less than 37·2ºC in the absence of antipyretic medication.” | 37.2 | 1 | 0 | 1 | 0 | 0 | 0 | 1 |
|  |  |  |  |  |  |  |  | 47 |

*Below is further description of each definition components:

Temperature cut-off: The different cut-off temperature to define fever in degree Celsius

Treatment administration: Mention of treatment or drug administration

Antibiotic administration: Mention of antibiotic administration

Antipyretic administration: Mention of antipyretic administration

24-hour window: Desired temperature maintained for 24 hours to declare fever clearance

48-hour window: Desired temperature maintained for 48 hours to declare fever clearance

Twice temperature measurement: Mention of two consecutive measurement in declaring fever clearance

0 and 1 symbolises ‘no’ and ‘yes’ respectively.
