## Supplementary material for "Systematic review of the scrub typhus treatment landscape: Assessing the feasibility of an individual participant-level data (IPD) platform": S5 Appendix. Variable dictionary

Research Questions

#### Primary Research Objectives

- What are the characteristics of scrub typhus treatment efficacy studies conducted in the last 20 years? The review will describe among other things treatment tested, patient characteristics, diagnostic methods, geographical location, outcome measures, and statistical methodology.

#### Secondary Research Objectives

- To assess the technical feasibility to develop a scrub typhus individual patient data platform.
- Is there scope for developing a global scrub typhus data platform for observational, diagnostic or prognostic data?

Objectives and Variables by Data Form

### 1. STUDY

#### Study identification and publication details

| **st_study_title** | Free text: Short Hand Reference (first author, publication year) and, *if applicable*, Scientific title of the study/trial, as stated at registration. E.g. ‘Botoni2007’ or ‘Morillo2017; STOPCHAGAS’ |
| --- | --- |
| **st_study_num** | Text box: Assign study number to the paper, use 3 digits e.g. 001, 025, 103  Define this as the secondary unique field to use it as secondary identifier and have it displayed across forms |
| **st_reg_id** | Free text: Trial registry identifier, if the trial was registered and registration could be found (i.e. mentioned in the publication, or identified through search in ClinicalTrials.gov, or WHO ICTRP registries). Enter “**-99**” if a registry ID is not found or unknown. Separate by semi colon if multiple registry identifiers  e.g. NCT9834808; ACT4576 |
| **st_primary_pub** | Free text: Reference of the journal article or conference abstract, including all authors and full title. Formatting style: ‘Vancouver’  *Author. Title. Journal. Year; Volume (Issue):Pages.* |
| **st_pub_type** | Dropdown variable: describing the medium with which research results were reported  1, “**Full text publication**” \| 2, “**Conference abstract**” \| 3, “**Trial registry**” \| 4, “**Dissertation/thesis**” \| 5, “**Other**”  NB: Only conference abstracts from 1 January 2016 or newer were analysed; it is assumed that studies presented at conference beforehand have been published by now and are therefore captured elsewhere. |
| **st_pub_date** | Date of publication (**DD/MM/YYYY**) (earliest known, i.e. chose ‘ePub ahead of print’, ‘Published online’ date if available), or of presentation at a conference. If an exact publication date is unknown (online or published version), if possible, chose ‘manuscript accepted’ date and add 1 month; estimate it from the journal; or, arbitrarily DD=15 and MM=01 and year of publication. |
| **st_author** | Free text: details of the author or contact for correspondence, i.e., name and institution |
| **st_author_contact** | Email address for the author or contact for correspondence |
| **st_oth_pub_no** | Number of additional publications (journal articles or conference abstracts) that provide supplementary information in addition to the primary study publication (i.e. protocol, subgroup analyses, sub-studies such as PK/PD results or follow-up results). If none enter “**0**”. |
| **st_oth_pub_1, 2, 3 etc** | Free text: Reference of the associated journal article or conference abstract, including all authors and full title. Formatting style: ‘Elsevier Harvard (with titles)’ |
| **st_shared** | Dropdown variable: Have the data sets related to this study been shared/submitted to IDDO? Discrete choice: 0, “**No**” \| 1, “**Yes**” \| 99, “**Unknown**” \| 3, “**Requested no response**”  As a default – all studies where data has yet to be requested - select “**Unknown**”.  If a dataset related to the study has been shared, including partial datasets – select “**Yes**”  If a request has been sent to a data contributor and the contributor has declined or refused to share the dataset – select “No”  If a request has been sent to a data contributor and no response has been received, nor has any dataset been submitted to IDDO – select “Requested no response” |
| **st_shared_iddo_id** | If **[st_shared] =** 1, Yes  Free text: What is the IDDO study ID assigned to the associated dataset? |
| **st_shared_notes** | If **[st_shared] =** 1, Yes  Free text: Details of datasets IDDO have received  i.e., all associated datasets, only molecular marker data, one of two cohorts/study sites etc. |

#### Cohort

##### Number of cohorts within single study

| **st_num_cohort** | Number of cohorts within this single ‘study’ reported in this article/abstract (see Appendix for details). |
| --- | --- |
| **st_coh_div_reas** | Checkbox variable: describing the Reason for the division of study cohorts: 1, “**Several countries**” \| 2, “**Different participant groups**” \| 3, “**Different protocols per group** ” |
| **st_coh_div_desc** | Free text: further justification for counting a single ‘study’ reported in this article/abstract as several ‘cohorts’ = set of participants sharing the same study meta-data characteristics (see Appendix for details). |

#### Setting

##### Where was the study conducted/cohort recruited?

##### The number of participants included in each cohort

| **Following variables completed for each cohort: 1,2,3 etc.** | |
| --- | --- |
| **st_coh1_country** | Dropdown variable: Name of the country of study. Country name based on UN classification: <http://unstats.un.org/unsd/methods/m49/m49regin.htm>  Taiwan and unknown options added. |
| **st_coh1_site** | Free text: most precise name and address of each and every site of recruitment, including longitude and latitude GPS coordinates, if available. |
| **st_coh1_region** | Dropdown variable: World region of the country of study. Use UN classification : <http://unstats.un.org/unsd/methods/m49/m49regin.htm> |
| **st_coh1_income** | Dropdown variable: describing the income group of the country; **“Low” “Lower middle” “Upper middle” “High”**: as per World Bank: <http://data.worldbank.org/country> |
| **st_coh1_site_cat** | Checkbox variable: describing the category of the site chosen as a recruitment centre of target participants: “**hospital/health-centre**”; “**community**” (village/city, households, etc.); “**other**”. Select “**unknown**” otherwise. |
| **st_coh1_setting** | Dropdown variable: describing the setting where the study was done: “**rural**”; “**semi-urban**”; “**urban**”. Use Google Earth if unclear in full text.  **Urban** if the area around the point, within 5x5 km square, is dominated by commercial buildings (e.g. offices, malls, apartments, high rise buildings).  **Suburban** if the area around the point, within 5x5 km square, is dominated by housing.  **Rural** if the area around the point, within 5x5 km square, is dominated by agricultural land/forest.  Several options may apply. Enter “**unknown**” otherwise. |
| **st_coh1_pt_setting** | Checkbox variable: describing the setting where recruited participants live: “**rural**”; “**semi-urban**”; “**urban**”. Several options may apply. If not mentioned or unclear, enter “**unknown**”. |
| **st_coh1_num_patients** | Integer: Total number of participants in the initial cohort; i.e. who were *INCLUDED* into the study *and received* the intended intervention (drug(s), control, placebo, no-treatment, etc.), with the intention to follow them up for outcome assessment. |
| **st_coh1_num_scrub** | Integer: Total number of participants with scrub typhus in the initial cohort; i.e. who were *INCLUDED* into the stud*y and received* the intended intervention (drug(s), control, placebo, no-treatment, etc.), with the intention to follow them up for outcome assessment. |
| **st_coh1_num_scrubfollow** | Integer: Number of relevant participants (i.e. among **st_coh1_num_scrub**) who were followed-up. If follow-up is detailed for each of the different time-points, enter the maximum number of participants followed-up. Enter “**-1**” if not applicable (e.g. retrospective studies or when ‘provide samples at follow-up’ is an inclusion criterion, meaning that the study cohort was defined *a posteriori*, based on compliance); and “**-99**” if unknown. |
| **st_coh1_num_calculation** | Free text: Details if patient number not reported and calculations that were performed for patient estimates. That is, the way in which the scrub typhus infected number of participants (**st_coh1_num_scrub**) or those followed-up (**st_coh1_num_scrubfollow)** was obtained from the total of included participants (**st_coh1_num_patients**), if applicable. |

#### Study Characteristics

##### What study design was used and was data collected prospectively or retrospectively?

##### Was the study comparative/were multiple arms involved in the study?

##### Were patients exposed to a treatment or intervention during the study and if so, how many arms did the study involve, including control-arms?

| **st_type** | Dropdown variable: detailing the study type. Discrete choice:  1, “**Interventional**” \| 2, “**Observational**”  *See Appendix for further description of categories* |
| --- | --- |
| **st_design** | Dropdown variable: detailing the study type/design. Discrete choice:  Dropdown variable: detailing the study design. Discrete choice:  If st_type is “**Interventional**” then select one of the following study designs.  1, “**RCT**” \| 2, “**Quasi-randomised**” \| 3, “**Non-randomised**”  Select 3 “non-randomised” for single arm interventional study arms.  If st_type is “**Observational”** then select one of the following study designs:  \| 4, “**Cohort-study**” \| 5, “**Case report**” \| 6 “**Case series”** \| 7, **“Case-control**” \| 8, “**Diagnostic test accuracy**” \| 9, “**Other**”  *See Appendix for further description of categories* |
| **me_phase** | If [**st_type**] = “1” 🡪 **Interventional**  In which clinical trial phase is this study?  Dropdown variable: 1, “**Phase 1**” \| 2, “**Phase 2**” \| 3, “**Phase 3**” \| 4, “**Phase 4**” \| 99, “**Unknown**”  Only select Phase if explicitly reported in the publication. DO not make any assumption or inference based on study design. |
| **st_pros_retro** | Dropdown variable: describing whether data collected for the study was performed prospective or retrospectively. Discrete choice:  0, “**Prospective**” \| 1, “**Retrospective**” |
| **st_comparative** | Dropdown variable: describing whether the study was comparative. Were all participants exposed to the same treatment/ intervention/ regimen or were multiple study arms compared. Discrete choice: 0, “**No**” \| 1, “**Yes**” |
| **st_intv_arms** | Integer: Number of study arms  Study arms: an arm/group of patients receiving a specific treatment regimen or intervention. I.e., also referred to as number of treatment arms, comparative arms, treatment groups, intervention groups |

##### Was assignment to different arms randomised and blinded?

| **st_random** | If [**st_type**] = “1” 🡪 **Interventional**  Dropdown variable: describing the level of randomisation and specification of methods reported. Discrete choice:  1, “**Single-arm**” \| 2, “**Multi-arm no randomisation reported**” \| 3, “**Multi-arm randomisation reported no detailed methods**” \| 4, “**Multi-arm randomisation reported with detailed methods**” \| 5, “**Other**” \| 99, “**Unknown**” |
| --- | --- |
| **me_random_desc** | If [**st_random**] = 4, “**Multi-arm randomisation reported with detailed methods**” or 5, “**Other**”  Free text: further details on randomisation methods. If single arm type 'single arm'. |
| **st_blind** | If [**st_type**] = “1” 🡪 **Interventional**  Dropdown variable: describing the level and methods of blinding reported. That is, for randomised studies, was it reported if the person delivering the drug and/or assessing the participant was blinded to the intervention administered? Discrete choice:  1, “**Single-arm**” \| 2, “**Multi-arm no blinding reported**” \|3, “**Multi-arm blinding reported no details who blinded**” \| 4, “**Multi-arm blinding reported with details who blinded**” \| 5, “**Other**” \| 99, “**Unknown**” |
| **me_blind­_desc** | If [**st_type**] = “1” 🡪 **Interventional**  If [**st_blind**] = 4, “**Multi-arm blinding reported with details who blinded**”  Free text: further details on the blinding methods  Including classification - single blind or double blind |
| **me_alloc_ratio** | If [**st_type**] = “1” 🡪 **Interventional**  Free text: describing how participants were allocated to each study arm e.g. 1:1, 1:2; 1:3, 1:4. If this is unknown, please type ‘Unknown’. If the study is single arm, please type ‘Single arm’. |
| **me_all_conc** | If [**st_type**] = “1” 🡪 **Interventional**  Dropdown variable: describing whether concealment was performed during participants allocation to each study arm. Discrete choice: 0, “**No**” \| 1, “**Yes**” \| 99, “**Unknown**” \| 3, “**Single arm**” |

##### Were sample size calculation and how it was done described?

| **me_samp_size** | Dropdown variable: describing whether sample size calculation was done. Discrete choice: 0, “**No**” \| 1, “**Yes**” \| 99, “**Unknown**” |
| --- | --- |
| **me_samp_size_desc** | If [**me_samp_size**] = 1, “**Yes**”  Free text: further details on sample size calculation |
| **me_samp_soft** | If [**me_samp_size**] = 1, “**Yes**”  Free text: describing the software used during sample size calculation e.g. PASS, SPSS, Stata, R, SAS, Epinfo, Minitab. If this is unknown, please type ‘Unknown. If other software was used, please type ‘Other’. |

##### When was the study conducted and for how long were patients followed-up?

| **st_start_date** | Start date of study (**DD/MM/YYYY**). If the exact date is unknown, then, arbitrarily, DD=15 and MM=01. If the year of study start date is unknown, refer to [**st_end_date]** and estimate the start year by subtracting the trial and follow-up duration from the [**st_end_date]** year. If follow-up duration is unknown, subtract one year from study end date. |
| --- | --- |
| **st_end_date** | End date of study (**DD/MM/YYYY**). If the exact date is unknown, if possible, estimate it from start date and duration of follow-up, or, arbitrarily DD=15 and MM=12. If the year of study end is unknown, arbitrarily select the year before the publication was submitted to or accepted for publication where provided or alternatively the year before the publication date. |
| **st_followup** | Follow-up duration in days. Longest reported endpoint / observation measured in any participant included in the study. Longest follow-up time point should be calculated with day 1 being day 1 of treatment initiation or if observational start of study (NOT number of days after treatment has concluded). If not explicitly stated but maximum length of hospitalisation/observation/symptoms observed for a patient is reported enter this time. If no maximum enter mean/median. Otherwise enter “**Unknown**.”  For calculations in days; assumptions 1 month=30 days, 1 year 365 days. Enter -99, |

##### Other relevant study characteristics

| **st_insecticidal** | Dropdown variable: describing the use of insecticidal or other measures to reduce reinfection during trial. Discrete choice: 0, “**No**” \| 1, “**Yes**” \| 99, “**Unknown**” |
| --- | --- |
| **st_strain** | Dropdown variable: describing whether strain type data was collected in the study. That is, was the specific strain of *Orientia tsutsugamushi* infection classified for each patient*.* Discrete choice: 0, “**No**” \| 1, “**Yes**” \| 99, “**Unknown**”  If PCR, IFA, IIP, or ELISA was used in the study at baseline but it is not specifically reported that analysis of the *O. tsutsugamushi* strains were performed, select “**Unknown**”. If only ICT was used in the study, select “**No**”. |
| **st_genotype** | Dropdown variable: describing whether genotypic data was collected in the study. Discrete choice: 0, “**No**” \| 1, “**Yes**” \| 99, “**Unknown**”  If PCR was used in the study at baseline but it is not specifically reported that genotyping of the *O. tsutsugamushi* strains were performed, select “**Unknown**”. If only serology was used in the study, select “**No**”. |
| **st_genotype_data** | If [**st_genotype**] = 1, “**yes**”  Free text: details of availability of genomic data e.g. publicly available on GenBank® database; Sanger Institute etc. Enter “**unknown**” if no database or access to data is mentioned in the article |

### 2. PARTICIPANTS

#### Diagnostic inclusion

##### What analyses were performed to diagnose scrub typhus?

| **pa_conf_dx** | Dropdown variable: was a confirmed scrub typhus diagnosis required for inclusion in the study? Discrete choice:  0, “**No**” \| 1, “**Yes**” \| 99, “**Unknown**”  *See Appendix for the definition of confirmed scrub typhus diagnosis* |
| --- | --- |
| **me_case** | Free text: Describe the case definition used for inclusion |
| **st_diag_meth** | Checkbox variable: describing the types of diagnostic tests used to confirm diagnosis of participants at baseline/study inclusion or during study if observational. Several options may apply:  1, “**STIC**” \| 2, “**Cell culture isolate**” \| 3, “**IFA**” \| 4, “**IIP**” \| 5, “**PCR**” \| 6, “**ELISA**” \| 7, “**ICT**” \| 8, “**Eschar presence**” \| 9, “**Weil-Felix**” \| 10, “**History and clinical manifestation**” \| 11, “**Other**” \| 99, “**Unknown**”  *See Appendix for further description of categories* |
| **st_diag_meth_oth** | If [**st_diag_meth**] = “11” 🡪 “**Other**”  Textbox: Mention other diagnostic methods used |
| **st_diag_blind** | Dropdown variable: Were diagnostic procedures blinded? Discrete choice: 0, “**No**” \| 1, “**Yes**” \| 99, “**Unknown**” |
| **st_diag_cci** | If [**st_diag_meth**] = “2” 🡪 **Cell culture isolate**  Checkbox variable: what sample was cultured?  Several options may apply :  1, “**Eschar**” \| 2, “**Blood**” \| 3, “**CSF**” \| 4, “**Other**” \| 99, “**Unknown**” |
| **st_diag_cci_details** | If [**st_diag_cci**] = “4” 🡪 **Other**  Free text: further details on other type of samples taken to culture *O. tsutsugamushi* |
| **st_diag_ifa** | If [**st_diag_meth**] = “3” 🡪 **IFA**  Checkbox variable: which antibody was targeted by the IFA?  Several options may apply :  1, “**IgM**” \| 2, “**IgG**” \| 3, “**IgA**” \| 99, “**Unknown**” |
| **st_diag_iip** | If [**st_diag_meth**] = “4” 🡪 **IIP**  Checkbox variable: which antibody was targeted by the IIP?  Several options may apply :  1, “**IgM**” \| 2, “**IgG**” \| 99, “**Unknown**” |
| **st_diag_ict** | If [**st_diag_meth**] = “7” 🡪 **ICT**  Checkbox variable: which antibody was targeted by the ICT?  Several options may apply :  1, “**IgM**” \| 2, “**IgG**” \| 3, “**IgA**” \| 99, “**Unknown**” |
| **st_diag_pcr_sample** | If [**st_diag_meth]** = “5” 🡪 **PCR**  Checkbox variable: which sample was used for DNA extraction and PCR?  Several options may apply :  1, “**Eschar**” \| 2, “**Blood**” \| 3, “**CSF**” \| 4, “**Other**” \| 99, “**Unknown**” |
| **st_diag_pcr_target** | If [**st_diag_meth]** = “5” 🡪 **PCR**  Checkbox variable: which target gene was amplified?  Several options may apply:  1, “**47 kDa**” \| 2, “**56 kDa**” \| 3, “**groEL**” \| 4, “**Other**” \| 99, “**Unknown**” |
| **st_diag_pcr_method** | If [**st_diag_meth**]= “5” 🡪 “**PCR**”  Checkbox variable: Were conventional PCR methods (i.e., looking for the presence of any genetic material; positive or negative results) or quantitative PCR methods (i.e., measurement of the amount of genetic material present) used? And was full genome sequencing performed. Several options may apply:  1, “**Conventional PCR**” \| 2, “**Real-time quantitative PCR**” \| 3, “**Nested PCR**” \|4, “**Full genome sequencing**” \| 99, “**Unknown**” |
| **st_strain_type** | If [**st_strain**] = “1” 🡪 “**Yes**”  Checkbox variable: Which strain(s) was/were included in the study? Several options may apply: 1, “**Karp**” \| 2, “**Kato**” \| 3, “**Gilliam**” \| 4, “**TA 716**” \| 5, ”**Boryong**” \| 6, “**TA 763**” \| 7, “**Saitama**” \| 8, “**Shimokoshi**” \| 99, “**Kawasaki**” \| 10, “**Kuroki**” \| 11, “**Other**” |
| **st_strain_type_oth** | If [**st_strain_type_oth] = “**11”, “**Other**”  Free text: What other strain(s) was/were included in the study? |
| **st_diag_meth_detail** | Free text: Further details of diagnostic method and technique used to confirm diagnosis and/or confirm eligibility for study inclusion. |

#### Other Inclusion Criteria

##### What is the age range and gender split of included patients?

| **st_elig_age_min** | Minimum age for eligibility in the study (inclusion criteria), that is the youngest age eligible for study participation, in years. If minimum is not reported, use lower limit of included participant age range. If no eligible age range reported for adult participants, arbitrarily enter “**18**”. |
| --- | --- |
| **st_elig_age_max** | Maximum age for eligibility in the study (inclusion criteria), that is the oldest age eligible for study participation, in years. If maximum is not reported, use upper limit of included participant age range. If no upper age range and no maximum age reported enter “**99**” |
| **pa_age_min** | Minimum age of included participants, that is the youngest participant included in the study, in years. If minimum is not reported, use lower limit of eligible age range. If no eligible age range reported for adult participants, arbitrarily enter “**18**”. Validation changed from integer to number to accommodate decimal numbers (i.e. if age is given in months). |
| **pa_age_max** | Maximum age of included participants, that is the eldest participant included in the study, in years. If maximum is not reported, use upper limit of eligible age range. If no upper age range and no maximum age reported enter “**99**”. Validation changed from integer to number to accommodate decimal numbers (i.e. if age is given in months). |
| **pa_age_mean** | Integer: The mean age of included participants in a study. Calculate if the mean is not available, but the age of all participants is known (e.g. described in the supplement). Enter “-99” if unknown. Validation changed from integer to number to accommodate decimal numbers. |
| **pa_age_median** | Integer: The median age of included participants in a study. Calculate if the median is not available, but the age of all participants is known (e.g. described in the supplement). Enter “-99” if unknown. Validation changed from integer to number to accommodate decimal numbers. |
| **pa_female** | Integer: Total number of female participants included in the study. If not reported and a ratio of M:F is provided, calculate the number of females by applying the ratio to the total number of included participants. Enter “**-99**” if unknown. |
| **pa_male** | Integer: Total number of male participant included in the study. If not reported and a ratio of M:F is provided, calculate the number of males by applying the ratio to the total number of included participants. Enter “**-99**” if unknown. |

##### Were immunocompromised patients or pregnant women included?

| **me_preg_yn** | Dropdown variable: were pregnant women included in the study? Discrete choice: 1, “**Included**” \| 2, “**Excluded**” \| 99, “**Unknown**”  Assumption made that pregnant women were “**Excluded**” in paediatric studies where age of participants was ≤10years. Enter “**Unknown**” otherwise. |
| --- | --- |
| **pa_preg_desc** | If [**me_preg_yn**] = 1, “**Included**”  Free text: further details on the eligibility criteria of pregnant participants |
| **me_hiv_yn** | Dropdown variable: were patients with HIV included in the study? Discrete choice:  1, “**Included**” \| 2, “**Excluded**” \| 99, “**Unknown**” |
| **pa_hiv_desc** | If [**me_hiv_yn**] = 1, “**Included**”  Free text: further details on the eligibility criteria of HIV+ participants |
| **me_imm_yn** | Dropdown variable: were immunocompromised patients (excluding HIV+) included in the study? Discrete choice:  1, “**Included**” \| 2, “**Excluded**” \| 99, “**Unknown**” |
| **pa_immuno_type** | If [**me_imm_yn]** = “1” 🡪 **Included**  Free text: type of immunocompromised condition (e.g. diabetes, tuberculosis, etc) |
| **me_mal_yn** | Dropdown variable: were malnourished patients included in the study? Discrete choice: 1, “**Included**” \| 2, “**Excluded**” \| 99, “**Unknown**” |
| **pa_malnut_cri** | If [**me_mal_yn**] = 1, “**Included**”  Dropdown variable: what criteria was used to diagnose malnutrition? Discrete choice: 1, **“Weight for height”**\| 2, **“Weight for age”** \| 3, **“Mid upper arm circumference”** \| 4, **“Triceps skinfold thickness”** \| 5, **“Body mass index”** \| 6, **“Others”** \| 99, **“Unknown”** |
| **pa_malnut_desc** | If [**me_mal_yn**] = 1, “**Included**”  Free text: further details on the eligibility criteria of malnourished participants. If [**pa_malnut_cri**] = 6, “**Others**”, mention the other criteria here. |

##### Other specific inclusion criteria

| **pa_protocol** | Free text: Assessment for ability and willingness to follow/minimal compliance to the protocol required for inclusion, especially in analyses (e.g. “able and willing to be examined by the study physician at baseline and follow-up”). |
| --- | --- |
| **pa_consent** | Free text: details/method for securing informed consent, including who gave consent for children (parent, head of the school, etc.) and how (orally, in writing, etc.). |
| **pa_oth_incl** | Free text: other inclusion criteria not falling into any of the previous categories. |

#### Exclusion criteria

| **pa_excl_scrub** | Free text: description of any scrub typhus specific criteria that were used to determine ineligibility/exclusion from the study. |
| --- | --- |
| **pa_excl_oth** | Free text: list of severe illnesses or systematic diseases/health conditions leading to exclusion. |
| **pa_excl_antb** | Dropdown variable: describing whether participants were excluded from the study if they had received any antibiotic treatment prior to study inclusion. Discrete choice: 0, “**No**” \| 1, “**Yes**” \| 99, “**Unknown**” |
| **pa_excl_othmed** | Free text: further details of other medication(s), including antibiotics taken during a specific period prior to (or during) the study that lead to exclusion. |
| **pa_oth_excl** | Free text: Other exclusion criteria not falling into any previous categories |

### 4. STUDY ARM

**NOTE: IN REDCap the ‘Study Arm’ Form is now a repeatable instrument that requires a new ‘study arm’ form to be created for each arm – This is not automatically created for you.**

**This replaces the old ‘Intervention’ form that would populate variables based on branch logic of the number of how many ‘treatment arms’ you entered.**

**The reason for these changes is to accurately capture instances where patients in a single study arm are exposed to multiple therapies/treatments/interventions.**

**The below variables need to be completed for each study arm in a separate ‘study arm’ form which is a repeatable instrument in REDCap.**

##### Details of the intervention/treatment regimens for each study arm and corresponding number of participants per study arm

| **Following variables completed for each treatment arm 1,2,3 etc.** | |
| --- | --- |
| **sa_target** | Dropdown variable: What was the treatments/interventions given? Discrete choice:  1, “**Antibiotic treatment**” \| 2, “**Symptomatic treatment**” \| 3, “**Other”** \| 99, “**Unknown**” |
| **sa_num_intv** | Integer: How many treatments/interventions were patients in this treatment arm exposed to?  Notes: To capture total number interventions in a single study arm – i.e., if combination therapy, multiple interventions or drug therapy plus another intervention(s)  For observational study, use this variable to capture antibiotic treatment administered for scrub typhus.  E.g. if combination treatment of drug A plus drug B was administered in a single study arm enter ‘2’ |
| **sa_combi** | If **[sa_num_intv]** 🡪 >=2  Dropdown variable: Were the multiple interventions administered in combination?  1, “**co-administered**” \| 2, “**sequential**” \| 3, **“other”**  Co-administered: Treatments are given to patients at or almost the same time e.g. concomitant, one after the other, on the same day, etc.  Sequential: Intervention exposure or treatments are administered in a sequence – e.g. drug A days 1-30, drug B days 31-60. |
| **sa_fixed** | If **[sa_combi] =** 1, “**co-administered**”  Dropdown variable: If the treatments were co-administered was it a fixed-dose formulation or loose-dose administration?  1, “**fixed-dose**” \| 2, “**loose-dose**” \| 99, “**unknown**”  Fixed-dose: defined as a combination drug is a fixed-dose combination (FDC) that includes two or more active pharmaceutical ingredients (APIs) combined in a single dosage form, which is manufactured and distributed in fixed doses.  Loose-dose: combination therapy that involves administration of two or more active pharmaceutical ingredients (APIs) in multiple dosage forms (i.e., **NOT** combined in a single dosage form) |
| **sa_num_tx** | Integer: Number of scrub typhus infected patients treated or exposed to the intervention. That is, the total number of scrub typhus patients in the study arm. |
| **sa_num_followup** | Integer: The reported number of patients followed-up in the study arm. If not reported, the maximum number of scrub typhus patients with at least one follow-up measurement post-baseline (i.e., during the treatment period or post-treatment follow-up) in the study arm.  Enter “-1” if not applicable (e.g. retrospective studies or when ‘provide samples at follow-up’ is an inclusion criterion, meaning that the study cohort was defined *a posteriori*, based on compliance); and “-99” if unknown. |
| *Branch logic – following variables to be completed for each intervention administered within the single study/treatment arm - corresponding to* **[sa_num_intv]** | |
| **sa_intv1_desc** | Free text: Name of treatment(s) or intervention specific to the study arm of interest. |
| **sa_intv1_drug** | If **[sa_num_intv]** >= 1  Dropdown variable: Name of drug administered / active pharmaceutical ingredient (API) as per INN. Discrete Choice  1, “**Doxycycline**” \|2, “**Minocycline**” \| 3, “**Chloramphenicol”** \| 4, “**Rifampicin**” \| 5, “**Azithromycin**” \| 6, “**Telithromycin**” \| 7, “**Levofloxacin**” \| 8, “**Other**” \| 99, “**Unknown**”  When the comparative arm is no treatment, select “**other**”.  *Controlled vocabulary: International Nonproprietary Names (INN) should be used to identify pharmaceutical substances or active pharmaceutical ingredients.*  If a fixed-dose combination (FDC) – in this variable select one of the APIs and the second API that makes up the single dosage form in the next variable [**in_intv1_drug_b]**. If there are more than 2 APIs in a single dose formulation – detail further in free text of **[in_intv1_reg_detail]**. |
| **sa_intv1_drug_oth** | If **[sa_intv1_drug]** = “8”, “**Other**”  Name of other medicine administered |
| **sa_intv1_reg_yn** | Dropdown: Are regimen details of the intervention or exposure available?  Discrete choice:  0, “**No**” \| 1, “**Yes**” |
| **sa_intv1_drug_b** | If **[sa_fixed]** = 1, “**fixed-dose**”  Free text: name of second drug/API that makes up the single dosage form. |
| **sa_intv1_route** | If [**sa_intv1_reg_yn**] = 1, yes  Dropdown variable: What was the route of administration?  1, “**Oral**” \| 2, “**Intravenous**” \| 3, “**Intramuscular**” \| 4, “**Intralymphatic**” \| 5, “**Subcutaneous**” \| 7, “**Other**” \| 99, “**Unknown**”  *EMA Controlled Vocabulary for Route of administrations: http://www.ema.europa.eu/docs/en_GB/document_library/Scientific_guideline/2009/09/WC500002730.pdf* |
| **sa_intv1_route_dur** | If **[sa_intv1_route]** = 2, “Intravenous”  Integer: Duration of infusion (in below units) |
| **sa_intv1_dur_un** | If **[sa_intv1_route_dur]** >= “1”, not “Unknown”  Dropdown variable: Unit of infusion duration time. Discrete choice:  1, “**Minutes**” \| 2, “**Hours**” \| 3, “**Days**” \| 4, “**Other**” \| 99, “**Unknown**” |
| **sa_intv1_route_other** | If **[sa_intv1_route]** =7, “Other”  Free text: Description of ‘Other’ route of administration |
| **sa_intv1_dose_method** | If [**sa_intv1_reg_yn**] = 1, yes  Dropdown variable: Method of dosing calculation. Discrete choice:  1, “**Target mg/kg**” \| 2, “**Weight-band**” \| 3, “**Age-band**” \| 4, “**other**” \| 99, “**unknown**” |
| **sa_intv1_other_det** | If [**sa_intv1_dose_method**] = 2, Weight-band, 3, Age-band, 4, Other  Free text: dose used per administration  Enter with unit |
| **sa_intv1_dose_mgkg** | If **[sa_intv1_dose_method]** = 1 “Target mg/kg”  Free text: Targeted dosing or range in mg/kg |
| **sa_intv1_freq_yn** | If [**sa_intv1_reg_yn**] = 1, yes  Dropdown: Are details of the frequency of administration of the drug(s)/treatment/intervention available?  Discrete choice:  0, “**No**” \| 1, “**Yes**” \| 99, “**Unknown**”  Note: If the frequency is reported as a range (eg. 1-2 times a day), select ‘no’. Enter any information provided to the ‘Detailed regimen’ box. |
| **sa_intv1_freq_con_yn** | If [**sa_intv1_freq_yn**] = 1, yes  Dropdown: Is the frequency of the regimen consistently applied ie., if there is a loading dose or changes in frequency over time, select ‘No’  Discrete choice: 0, “**No**” \| 1, “**Yes**” |
| **~~sa_intv1_freq_discrete~~** | ~~Dropdown variable: frequency that the drug(s)/treatment/intervention was administered~~  ~~1, “~~**~~Once Daily~~**~~” \| 2, “~~**~~Twice Daily~~**~~” \| 3, “~~**~~Three times daily~~**~~” \| 4 “~~**~~Alternate Days~~**~~” \| 5, “~~**~~Other~~**~~” \| 99, “~~**~~Unknown~~**~~”~~  @HIDDEN added in field annotation |
| **sa_intv1_freq** | If [**sa_intv1_freq_yn**] = 1, yes  Integer: frequency that the drug(s)/treatment/intervention was administered for a given time period  -99 if unknown  e.g. Twice daily (Frequency = 2; Time period units = days; time period magnitude = 1)  e.g. Once every two days (Frequency = 1; time period units = days; time period magnitude = 2)  e.g. Once every three months (Frequency = 1; time period units = months; time period magnitude = 3) |
| **sa_intv1_freq_time_un** | If [**sa_intv1_freq_yn**] = 1, yes  Dropdown variable: Units of given time period  1, “**Hours**” \| 2, “**Days**” \| 3, “**Weeks**” \| 4 “**Months**” \| 5, “**Years**” \| 6, “**Other**” 99, “**Unknown**”  e.g. Twice daily (Frequency = 2; Time period units = days; time period magnitude = 1)  e.g. Once every two days (Frequency = 1; time period units = days; time period magnitude = 2)  e.g. Once every three months (Frequency = 1; time period units = months; time period magnitude = 3) |
| **sa_intv1_freq_time_un_oth** | If [**sa_intv1_freq_time_un] = 6, other**  If [**sa_intv1_freq_time_un**] = 6, other  **Free text: Other unit of time.** |
| **sa_intv1_freq_time_mag** | If [**sa_intv1_freq_yn**] = 1, yes  Integer: Magnitude of given time period  -99 if unknown  e.g. Twice daily (Frequency = 2; Time period units = days; time period magnitude = 1)  e.g. Once every two days (Frequency = 1; time period units = days; time period magnitude = 2)  e.g. Once every three months (Frequency = 1; time period units = months; time period magnitude = 3) |
| **~~sa_intv1_dura~~** | ~~Integer: Duration in days of the length of treatment or days the intervention was administered. Enter “~~**~~-99~~**~~” if unknown. If duration of treatment varied, enter maximum number of days any included participant received the intervention. For calculations in days; assumptions 1 month=30 days, 1 year 365 days.~~  @HIDDEN added in field annotation |
| **sa_intv1_dura_fix_mag** | If [**sa_intv1_reg_yn**] = 1, yes  Integer: Duration of the **intended** (in the protocol or in guidelines if applicable) length of treatment or time the intervention was administered. Enter “**-99**” if unknown. If a range is given i.e., 7-14 days, please enter “**-1**” and provide the details in free text for the variable below **sa_intv1_reg_detail** |
| **sa_intv1_dura_fix_un** | If [**sa_intv1_dura_fix_mag**] **>**= “1”, not “Unknown”  Dropdown variable: Units for duration. Discrete choice: **1, Days \| 2, Weeks \| 3, Months \| 4, Years \| 9, Other** |
| **sa_intv1_dura_fix_un_oth** | If [**sa_intv1_dura_fix_un**] = 9, Other  Free text: Other units for duration |
| **sa_intv1_reg_detail** | Free text: Further details and description of the treatment or intervention regimen specific to the study arm of interest, including the dose description and details of frequency  Include details of how multiple therapies administered if **[In_intv1_combi]** = 3 “**other**” |
| **sa_intv1_supervision** | Dropdown variable: Was treatment administration supervised? Discrete choice:  0, “**No**” \| 1, “**Yes**” \| 99, “**Unknown**”  No: It is reported the treatment was not supervised  Yes: It is reported the treatment was supervised or administration/intervention delivered in a hospital/health centre setting  Unknown: Not-reported |

### 5. OUTCOMES

#### Primary Treatment Related Outcomes

##### Were treatment/intervention related outcomes assessed at any time during the study post-baseline (during the treatment period or follow-up)?

| **ou_assessed** | Dropdown variable: Were treatment/intervention related outcomes assessed at any time during the study post-baseline (during the treatment period or follow-up)? Discrete choice: 0, “**No**” \| 1, “**Yes**” |
| --- | --- |

##### How was treatment response expressed as an outcome, how was it measured and how often was it measured?

| **ou_fevclr** | Dropdown variable: Were fever clearance assessed at any time during the study post-baseline (during the treatment period or follow-up)?  Discrete choice: 0, “**No**” \| 1, “**Yes**” \| 99, “**Unknown**” |
| --- | --- |
| **ou_fevclr_measure** | If **[ou_fevclr]** = 1, “Yes”  Checkbox variable: How were fever clearance measured? Several options may apply: 1, “**Oral**” \| 2, “**Axillary**” \| 3, “**Core**” \| 4, “**Aural**” \| 99, “**Unknown”** |
| **ou_fevclr_def** | If **[ou_fevclr]** = 1, “Yes”  Free text: description of how the fever clearance was expressed i.e. definition of fever clearance. |
| **ou_fevclr_endpoints** | If **[ou_fevclr]** = 1, “Yes”  Integer: How often were participants followed-up for fever clearance assessments at any time during the study post-baseline (during the treatment period or follow-up), i.e. total number of fever clearance related measurements. Enter “**-99**” if unknown. |
| **ou_fevclr_length** | If **[ou_fevclr]** = 1, “Yes”  Integer: the time in days, for fever clearance follow-up endpoint, counted from day of first treatment/intervention/start of study. Enter “**-99**” if unknown. For calculations in days; assumptions 1 month=30 days, 1 year 365 days. |
| **ou_fevclr_time** | Dropdown variable: Were fever clearance time/time to defervescence assessed at any time during the study post-baseline (after treatment was given or during follow-up period)?  Discrete choice: 0, “**No**” \| 1, “**Yes**” \| 99, “**Unknown**” |
| **ou_fevclr_time_details** | If **[ou_fevclr_time]** = 1, “Yes”  Free text: Further details on fever clearance time measurements. |
| **ou_mort** | Dropdown variable: Were mortality assessed at any time during the study post-baseline (during the treatment period or follow-up)?  Discrete choice: 0, “**No**” \| 1, “**Yes**” \| 99, “**Unknown**”  Mortality is considered to be assessed if: (1) it is explicitly mentioned i.e. mortality is an outcome of interest that will be specifically recorded), (2) mortality reported in the results, (3) adverse events were recorded. |
| **ou_mort_length** | If **[ou_mort]** = 1, “Yes”  Integer: the time in days, for mortality follow-up endpoint, counted from day of first treatment/intervention/start of study. Enter “**-99**” if unknown. For calculations in days; assumptions 1 month=30 days, 1 year 365 days. |
| **ou_clinres** | Dropdown variable: Were clinical response/cure/recovery assessed at any time during the study post-baseline (during the treatment period or follow-up)?  Discrete choice: 0, “**No**” \| 1, “**Yes**” \| 99, “**Unknown**” |
| **ou_clinres_feverclr** | If **[ou_ clinres]** = 1, “Yes”  Dropdown variable: Were clinical response/cure defined as fever clearance?  Discrete choice: 0, “**No**” \| 1, “**Yes**” \| 99, “**Unknown**” |
| **ou_clinres_def** | If **[ou_ clinres_feverclr]** = 0, “No”  Free text: description of how clinical response to treatment was expressed i.e. definition of clinical response. |
| **ou_clinres_endpoints** | If **[ou_ clinres_feverclr]** = 0, “No”  Integer: How often were participants followed-up for clinical response/cure at any time during the study post-baseline (during the treatment period or follow-up), i.e. total number of fever clearance related measurements. Enter “**-99**” if unknown. |
| **ou_clinres_length** | If **[ou_ clinres_feverclr]** = 0, “No”  Integer: the time in days, for clinical response/cure follow-up endpoint, counted from day of first treatment/intervention/start of study. Enter “**-99**” if unknown. For calculations in days; assumptions 1 month=30 days, 1 year 365 days. |
| **ou_scrcomp** | Dropdown variable: Were scrub typhus complications assessed at any time during the study post-baseline (during the treatment period or follow-up)?  Discrete choice: 0, “**No**” \| 1, “**Yes**” \| 99, “**Unknown**” |
| **ou_scrcomp_measure** | If **[ou_scrcomp]** = 1, “Yes”  Checkbox variable: How were scrub typhus complication measured during follow up? |
| **ou_scrcomp_def** | If **[ou_scrcomp]** = 1, “Yes”  Free text: description of how scrub typhus complications were expressed i.e. definition of complications. |
| **ou_scrcomp_type** | If **[ou_scrcomp]** = 1, “Yes”  Checkbox variable: What type(s) of scrub typhus complications were noted?  1, “**Respiratory**” \| 2, “**Neurological**” \| 3, “**Haematological**” \| 4 “**Hepatological**” \| 5, “**Nephrological**” \| 6, “**Other**” \| 99, “**Unknown**” |
| **ou_scrcomp_type_other** | If **[ou_scrcomp_type]** = 6, “Other”  Free text: Description of other types of scrub typhus complications |
| **ou_scrcomp_type_details** | If **[ou_scrcomp]** = 1, “Yes”  Free text: Further description of diagnosis and conditions counted as scrub typhus complications e.g. noting acute respiratory disease syndrome (ARDS) if **[ou_scrcomp_type]** 🡪 1, “Respiratory”. |
| **ou_ scrcomp _endpoints** | If **[ou_ scrcomp]** = 1, “Yes”  Integer: How often were participants followed-up for scrub typhus complications assessments at any time during the study post-baseline (during the treatment period or follow-up), i.e. total number of fever clearance related measurements. Enter “**-99**” if unknown. |
| **ou_ scrcomp _length** | If **[ou_ scrcomp]** = 1, “Yes”  Integer: the time in days, for scrub typhus complications follow-up endpoint, counted from day of first treatment/intervention/start of study. Enter “**-99**” if unknown. For calculations in days; assumptions 1 month=30 days, 1 year 365 days. |
| **ou_labp** | Dropdown variable: Were improvement of laboratory parameters assessed at any time during the study post-baseline (during the treatment period or follow-up)?  Discrete choice: 0, “**No**” \| 1, “**Yes**” \| 99, “**Unknown**” |
| **ou_labp_measure** | If **[ou_labp]** = 1, “Yes”  Checkbox variable: What laboratory parameters were measured during follow up? Several options may apply: 1, “**Haematological**” \| 2, “**Liver function**” \| 3, “**Kidney function**” \| 4, “**Inflammatory markers**” \| 5, “**Other**” \| 99, “**Unknown”** |
| **ou_labp_details** | If **[ou_labp]** = 1, “Yes”  Free text: Further description of what laboratory parameters were tested. |
| **ou_labp_endpoints** | If **[ou_labp]** = 1, “Yes”  Integer: How often were participants followed-up for laboratory parameters at any time during the study post-baseline (during the treatment period or follow-up), i.e. total number of fever clearance related measurements. Enter “**-99**” if unknown. |
| **ou_labp_length** | If **[ou_labp]** = 1, “Yes”  Integer: the time in days, for laboratory parameters improvement, counted from day of first treatment/intervention/start of study. Enter “**-99**” if unknown. For calculations in days; assumptions 1 month=30 days, 1 year 365 days. |
| **ou_endpoint_details** | Free text: Further details or clarification of follow-up measurements |

#### Safety and Other Patient Related Outcomes

##### Were adverse events / side effects / safety variables assessed any time during the study post-baseline (during the treatment period or follow-up)?

##### Details of other patient related outcome measurements not captured above

| **ou_ae** | Dropdown variable: Were adverse events / side effects / safety variables assessed any time during the study post-baseline (during the treatment period or follow-up)? Discrete choice: 0, “**No**” \| 1, “**Yes**” |
| --- | --- |
| **ou_ae_type** | If [**ou_ae**] = 1, “Yes”  Dropdown variable: Were adverse events assessed consisted of mortality and/or complications only?  0, “**No**” \| 1, “**Yes, mortality only**” \| 2, “**Yes, complication only**” \| 3, “**Yes, mortality and complication**” |
| **ou_pt_outcome** | Checkbox variable: Were any of the following outcome measures assessed at any time during the study post-baseline (during the treatment period or follow-up)? Several options may apply:  1, “**Relapse**” \| 2, “**Hospital stay**” \| 3, “**Imaging parameters improvement**” \| 4, “**Pharmacokinetics**” |
| **ou_pt_outcome_details** | Free text: Further details on the above outcome measures, including: definition, how it was measured. |
| **ou_other_outcome** | Free text: Details of any other patient related outcomes measured/assessed at any time during the study post-baseline (during the treatment period or follow-up) that are not captured in above categories. |

#### In-Vitro Outcomes

##### Were in-vitro data reported?

| **me_iv_yn** | Dropdown variable: Were in vitro data reported? Discrete choice: 0, “**No**” \| 1, “**Yes**” |
| --- | --- |
| **me_iv_desc** | If [**me_iv_yn**] = 1, “Yes”  Free text: further details on in-vitro data reported |

#### Outcomes classification

##### Which variable is the primary endpoint?

##### How many other endpoints does this study evaluate?

| **me_prim_end** | Dropdown variable: the name of primary endpoint  Discrete choice: 1, **Mortality/death** \| 2, **Cure/recovery** \| 3, **Failure** \| 4, **Fever clearance/defervescence** \| 5, **Fever clearance time/time to defervescence** \| 6, **Complications** \| 7, **Relapse** \| 8, **Length of hospital stay** \| 9, **Improved laboratory parameters** \| 10, **Adverse events** \| 11, **Other** |
| --- | --- |
| **me_prim_end_oth** | If [**me_prim_end**] = 11, “**Other**”  Free text: further details on other name of primary endpoint |
| **me_prim_end_def** | Free text: the definition of primary endpoint |
| **me_prim_end_time** | Integer: the time point for primary endpoint (in days) |
| **me_num_end** | Dropdown list: 1, “**1**” \| 2, “**2**” \| 3, “**3**” \| 4, “**4**” \| 5, “**5**” \| 6, “**6**” \| 7, “**0**”  Number of other endpoint(s) |
| **me_end1** | If [**me_num_end**] >= “**1**”  Dropdown variable: name of other endpoint - 1  Discrete choice: Discrete choice: 1, **Mortality/death** \| 2, **Cure/recovery** \| 3, **Failure** \| 4, **Fever clearance/defervescence** \| 5, **Fever clearance time/time to defervescence** \| 6, **Complications** \| 7, **Relapse** \| 8, **Length of hospital stay** \| 9, **Improved laboratory parameters** \| 10, **Adverse events** \| 11, **Other** |
| **me_end1_oth** | If [**me_end1**] = 11, “**Other**”  Free text: further details on other name of other endpoint |
| **me_end1_def** | If [**me_num_end**] >= “**1**”  Free text: definition of other endpoint - 1 |
| **me_end2** | If [**me_num_end**] >= “**2**”  Dropdown variable: name of other endpoint - 2  Discrete choice: Discrete choice: 1, **Mortality/death** \| 2, **Cure/recovery** \| 3, **Failure** \| 4, **Fever clearance/defervescence** \| 5, **Fever clearance time/time to defervescence** \| 6, **Complications** \| 7, **Relapse** \| 8, **Length of hospital stay** \| 9, **Improved laboratory parameters** \| 10, **Adverse events** \| 11, **Other** |
| **me_end2_oth** | If [**me_end2**] = 11, “**Other**”  Free text: further details on other name of other endpoint |
| **me_end2_def** | If [**me_num_end**] >= “**2**”  Free text: definition of other endpoint - 2 |
| **me_end3** | If [**me_num_end**] >= “**3**”  Dropdown variable: name of other endpoint - 3  Discrete choice: Discrete choice: 1, **Mortality/death** \| 2, **Cure/recovery** \| 3, **Failure** \| 4, **Fever clearance/defervescence** \| 5, **Fever clearance time/time to defervescence** \| 6, **Complications** \| 7, **Relapse** \| 8, **Length of hospital stay** \| 9, **Improved laboratory parameters** \| 10, **Adverse events** \| 11, **Other** |
| **me_end3_oth** | If [**me_end3**] = 11, “**Other**”  Free text: further details on other name of other endpoint |
| **me_end3_def** | If [**me_num_end**] >= “**3**”  Free text: definition of other endpoint - 3 |
| **me_end4** | If [**me_num_end**] >= “**4**”  Dropdown variable: name of other endpoint - 4  Discrete choice: Discrete choice: 1, **Mortality/death** \| 2, **Cure/recovery** \| 3, **Failure** \| 4, **Fever clearance/defervescence** \| 5, **Fever clearance time/time to defervescence** \| 6, **Complications** \| 7, **Relapse** \| 8, **Length of hospital stay** \| 9, **Improved laboratory parameters** \| 10, **Adverse events** \| 11, **Other** |
| **me_end4_oth** | If [**me_end4**] = 11, “**Other**”  Free text: further details on other name of other endpoint |
| **me_end4_def** | If [**me_num_end**] >= “**4**”  Free text: definition of other endpoint - 4 |
| **me_end5** | If [**me_num_end**] >= “**5**”  Dropdown variable: name of other endpoint - 5  Discrete choice: Discrete choice: 1, **Mortality/death** \| 2, **Cure/recovery** \| 3, **Failure** \| 4, **Fever clearance/defervescence** \| 5, **Fever clearance time/time to defervescence** \| 6, **Complications** \| 7, **Relapse** \| 8, **Length of hospital stay** \| 9, **Improved laboratory parameters** \| 10, **Adverse events** \| 11, **Other** |
| **me_end5_oth** | If [**me_end5**] = 11, “**Other**”  Free text: further details on other name of other endpoint |
| **me_end5_def** | If [**me_num_end**] >= “**5**”  Free text: definition of other endpoint - 5 |
| **me_end6** | If [**me_num_end**] >= “**6**”  Dropdown variable: name of other endpoint - 6  Discrete choice: Discrete choice: 1, **Mortality/death** \| 2, **Cure/recovery** \| 3, **Failure** \| 4, **Fever clearance/defervescence** \| 5, **Fever clearance time/time to defervescence** \| 6, **Complications** \| 7, **Relapse** \| 8, **Length of hospital stay** \| 9, **Improved laboratory parameters** \| 10, **Adverse events** \| 11, **Other** |
| **me_end6_oth** | If [**me_end6**] = 11, “**Other**”  Free text: further details on other name of other endpoint |
| **me_end6_def** | If [**me_num_end**] >= “6”  Free text: definition of other endpoint - 6 |

### 6. ANALYSIS

#### Lost to Follow-Up (LFU)

##### Was LFU reported?

##### How was LFU defined and how was LFU handled in the analysis?

| **me_lfu_yn** | Dropdown variable: Were LFU reported? Discrete choice: 0, “**No**” \| 1, “**Yes**” \| 99, “**Unknown**” |
| --- | --- |
| **me_lfu_def** | If [**me_lfu_yn**] = 1, “**Yes**”  Free text: Further details on the definition of LFU |
| **me_lfu_analysis** | If [**me_lfu_yn**] = 1, “**Yes**”  Dropdown variable: How were LFU handled in the analysis? Discrete choice: 1, “**Excluded**” \| 2, “**Censored**” \| 3, “**Imputed**” \| 4, “**Other**” \| 99, “**Unknown**” |
| **me_lfu_analysis_desc** | If [**me_lfu_yn**] = 1, “**Yes**”  Free text: further description of how LFU was handled in the analysis |

#### Data Analysis

##### What analysis methods were used for endpoints

##### Was interim analysis carried out?

##### Details on Data Safety and Monitoring Board (DSMB) and software used for data analysis

| **me_prim_analysis** | Free text: Further details on the analysis method for primary endpoint |
| --- | --- |
| **me_end1_anlysis** | If [**ou_other_num**] = 1, “**1**”  Free text: Further details on the analysis method for other endpoint 1 |
| **me_end2_anlysis** | If [**ou_other_num**] = 2, “**2**”  Free text: Further details on the analysis method for other endpoint 2 |
| **me_end3_anlysis** | If [**ou_other_num**] = 3, “**3**”  Free text: Further details on the analysis method for other endpoint 3 |
| **me_end4_anlysis** | If [**ou_other_num**] = 4, “**4**”  Free text: Further details on the analysis method for other endpoint 4 |
| **me_end5_anlysis** | If [**ou_other_num**] = 5, “**5**”  Free text: Further details on the analysis method for other endpoint 5 |
| **me_interim_yn** | Dropdown variable: Was interim analysis carried out? Discrete choice: 0, “**No**” \| 1, “**Yes**” \| 99, “**Unknown**” |
| **me_interim_desc** | If [**me_interim_yn**] = 1, “**Yes**”  Free text: Further details on interim analysis |
| **me_dsmb_yn** | Dropdown variable: Was there any mention of Data Safety and Monitoring Board (DSMB)? Discrete choice: 0, “**No**” \| 1, “**Yes**” \| 99, “**Unknown**” |
| **me_dsmb_desc** | If [**me_dsmb_yn**] = 1, “**Yes**”  Free text: Further description on DSMB |
| **me_analysis_soft** | Dropdown variable: describing the software used for data analysis. Discrete choice: **1, Excel \| 2, SPSS \| 3, Stata \| 4, R \| 5, SAS \| 6, Epinfo \| 7, Minitab \| 99, Unknown \| 9, Other** |
| **me_analysis_soft_oth** | If [**me_analysis_soft**] = "9", “**Other**”  Free text: Software used for the data analysis - specify other |

### 7. REPORTING

#### Ethics

##### Was ethical approval reported?

##### Details on informed consent

| **me_ethic_yn** | Dropdown variable: Was ethical approval mentioned? Discrete choice: 0, “**No**” \| 1, “**Yes**” \| 2, “**Unknown**” |
| --- | --- |
| **me_ethic_desc** | If [**me_ethic_yn**] = 1, “**Yes**”  Free text: Further details on ethical approval |
| **me_consent_yn** | Dropdown variable: Was informed consent taken before enrolment? Discrete choice: 0, “**No**” \| 1, “**Yes**” \| 99, “**Unknown**” |

#### Funding and conflict of interest

##### Was funding source reported?

##### Was any conflict of interest reported?

##### Further details on funding and conflict of interest

| **me_fund** | Free text: Please copy the text describing funding from the publication. |
| --- | --- |
| **me_fund_type** | Checkbox variable: From what type of organisation did the funding come from? Several options may apply: 1, “**Government**” \| 2, “**International organisation**” \|3, “**University grant**” \| 4, “**Pharma company**” \| 5, “**Other**” \| 6, “**Not applicable (if no funding sources)**” \| 99, “**Unknown**” |
| **me_fund_type_gov_desc** | If [**me_fund_type**] = 1, “**Government**”  Free text: The name of governmental funding source |
| **me_fund_type_int_desc** | If [**me_fund_type**] = 2, “**International organisation**”  Free text: The name of international organisation funding source |
| **me_fund_type_uni_desc** | If [**me_fund_type**] = 3, “**University grant**”  Free text: The name of university grant funding source |
| **me_fund_type_pha_desc** | If [**me_fund_type**] = 4, “**Pharma company**”  Free text: The name of pharmaceutical company funding source |
| **rp_fund_name_oth** | If [**me_fund_type**] = 5, “**Other**”  Free text: The name of other funding source |
| **me_coi_yn** | Dropdown variable: Was any conflict of interest reported by the authors? Discrete choice: 0, “**No**” \| 1, “**Yes**” \| 99, “**Unknown**” |
| **me_coi_desc** | If [**me_coi_yn**] = 1, “Yes”  Free text: Please copy the text describing the conflict of interest |
| **me_coi_num** | If [**me_coi_yn**] = 1, “Yes”  Integer: Number of authors declaring conflict of interest |
| **me_coi_type** | If [**me_coi_yn**] = 1, “Yes”  Checkbox variable: further details on the type of conflict of interest reported. Several options may apply: 1, “**Funding from pharma manufacturing study drug to attend conferences**” \| 2, “**Funding from pharma manufacturing study drug for current or other studies**” \| 3, “**Works for pharma funding the trial**” 4, \| “**Works for other agency funding this study**” \|5, “**Other**” \| 99, “**Unknown**” |
| **me_coi_type_oth** | If [**me_coi_type(5)**] = '1', Other  Free text: What was the COI type? - specify other |

Appendix

#### Reasons for considering a single study as several cohorts

A study is to be divided accordingly into different cohorts if and only if at least one of the three following conditions apply:

1. Multi-centric study, with centres in several countries → 1 cohort per country
2. The study clearly differentiates a priori between different categories of participants → 1 cohort per participant group
3. The protocol is otherwise amended depending on site/group (e.g. different diagnostic approach, different follow-up time-points) → 1 cohort per protocol

#### Detailed explanation of study type and definitions

Use the following clinicaltrials.gov definitions of study type:

**Interventional (experimental) Study:** A type of clinical study in which participants are assigned to groups that receive one or more intervention/treatment (or no intervention) so that researchers can evaluate the effects of the interventions on biomedical or health-related outcomes. The assignments are determined by the study's protocol. Participants may receive diagnostic, therapeutic, or other types of interventions.

**Observational Study:** The general design of the strategy for identifying and following up with participants during an observational study. Types of observational study models include cohort, case-control, case-only, case-cross-over, ecologic or community studies, family-based, and other.

#### Detailed explanation of study design categories

Definitions of study designs 1-5 are as per the Cochrane Consumers & Communication Review Group

Study Design Guide. For Review Authors. June 2013. <https://cccrg.cochrane.org/sites/cccrg.cochrane.org/files/public/uploads/Study_design_guide2013.pdf>

Definitions of study design 6, Diagnostic Test Accuracy is as per The Joanna Briggs Institute Reviewers’ Manual 2015. The systematic review of studies of diagnostic test accuracy. <https://joannabriggs.org/assets/docs/sumari/Reviewers-Manual_The-systematic-review-of-studies-of-diagnostic-test-accuracy.pdf>

**Interventional (experimental) Study Designs**

1, “**RCT**”

RANDOMISED CONTROLLED TRIALS

In RCTs the investigator randomly assigns people to groups that will receive (intervention group) or not receive (control group) one or more interventions. The outcomes measured are then compared between the groups.

2, “**Quasi-randomised**”

QUASI-RANDOMISED CONTROLLED TRIALS

Trials that attempt to randomly assign participants to groups but use an inadequate approach to generate the random sequence are designated as quasi-randomised controlled trials. Such trials do attempt to randomly allocate participants with the intent of producing equivalent groups, but the randomisation methods used are not adequate because in practice they are relatively easy to manipulate or predict. Inadequate randomisation approaches: Alternation, Case record numbers, Birth dates, Week days or month of the year.

3, “**Non-randomised**”

Either a single arm interventional study or a multi-arm study that does not apply randomisation when assigning individuals to treatment.

**Observational Study Designs**

4, “**Cohort-study**”

COHORT STUDY (SYNONYM: FOLLOW-UP, INCIDENCE, LONGITUDINAL, PROSPECTIVE STUDY)

An observational study in which a defined group of people (the cohort) is followed over time. This also includes non-comparative studies e.g. “prospective observational” that followed only one cohort. The outcomes of people in subsets of this cohort are compared, to examine for example people who were exposed or not exposed (or exposed at different levels) to a particular intervention or other factor of interest. A cohort can be assembled in the present and followed into the future (this would be a prospective study or a "concurrent cohort study"), or the cohort could be identified from past records and followed from the time of those records to the present (this would be a retrospective study or a "historical cohort study"). Because random allocation is not used, matching or statistical adjustment at the analysis stage must be used to minimise the influence of factors other than the intervention or factor of interest.

5, “**Case-control**”

CASE-CONTROL STUDY (SYNONYMS: CASE REFERENT STUDY, RETROSPECTIVE STUDY)

A study that starts with identification of people with the disease or outcome of interest (cases) and a suitable control group without the disease or outcome. The relationship of an attribute (intervention, exposure or risk factor) to the outcome of interest is examined by comparing the frequency or level of the attribute in the cases and controls. For example, to determine whether thalidomide caused birth defects, a group of children with birth defects (cases) could be compared to a group of children without birth defects (controls). The groups would then be compared with respect to the proportion exposed to thalidomide through their mothers taking the tablets. Case-control studies are sometimes described as being retrospective as they are always performed looking back in time.

6 & 7, “**Case-report or series**”

CASE STUDY (SYNONYMS: ANECDOTE, CASE HISTORY, SINGLE CASE REPORT)

An uncontrolled observational study involving an intervention and outcome for a single person (or other unit).

CASE SERIES

An uncontrolled observational study involving an intervention and outcome for more than one person.

8, “**Diagnostic test accuracy**”

Diagnostic test accuracy studies compare a diagnostic test of interest (the ‘index test’) to an existing diagnostic test (the ‘reference test’), which is known to be the best test currently available for accurately identifying the presence or absence of the condition of interest. The outcomes of the two tests are then compared with one another in order to evaluate the accuracy of the index test. There are two main types of studies of DTA. The first is the diagnostic case-control design, also sometimes called the ‘two gate design’. In this study design people with the condition (cases) come from one population (i.e. a health care centre for people known to have the condition), while people without the condition come from another. Although this design gives an indication of the maximum accuracy of the test, the results will generally give an exaggerated indication of the test’s accuracy in practice. The second study design is cross-sectional, and involves all patients suspected of having the condition of interest undergoing the index test and the reference test. Those who test positive for the condition by the reference test can be considered to be the cases, whereas those who test negative are the controls.

9, “**Other**” Any other study design not captured in categories 1-8.

#### Classification of clinical studies

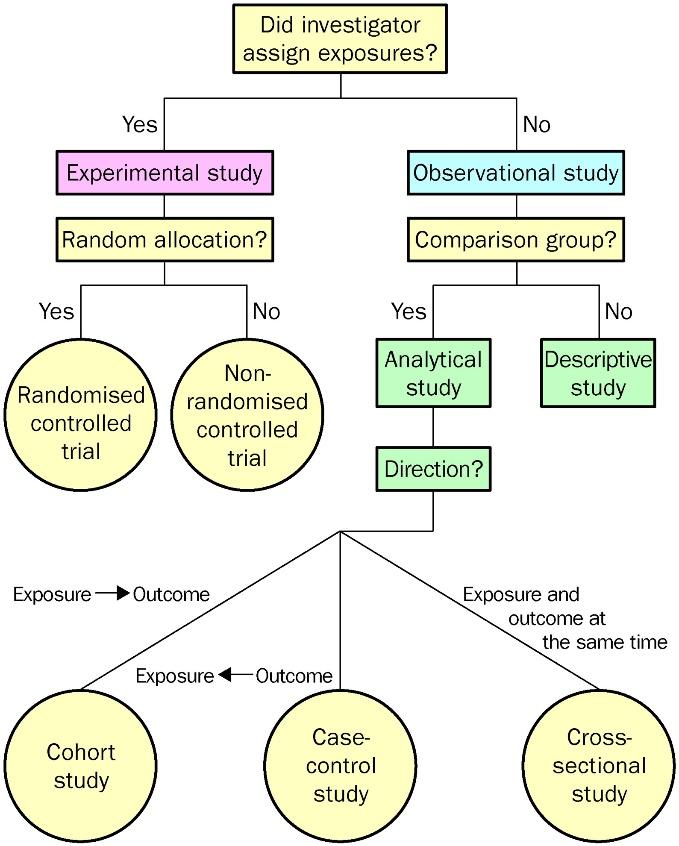

Figure 1, from Grimes DA, Schulz KF. An overview of clinical research: the lay of the land. *The Lancet* 2002;**359**:57–61. doi:[10.1016/S0140-6736(02)07283-5](https://doi.org/10.1016/S0140-6736(02)07283-5)

#### Definition of confirmed scrub typhus diagnosis

Definition of confirmed cases: history (i.e. went to endemic area or known foci of infection), clinical signs and symptoms (fever and other flu-like symptoms, eschar, organ dysfunction), and laboratory confirmation (PCR/IFA/IIP/ELISA/ICT/Weil-Felix).

#### Detailed explanation of diagnostic methods

1, “**STIC**”

SCRUB TYPHUS INFECTION CRITERIA (STIC) have been proposed as a more robust method to diagnose scrub typhus with more confidence, by including a panel of parameters with high specificity: (i) isolation of *O. tsutsugamushi*, (ii) at least two positive out of the three PCR assays targeting the 56kDa, 47kDa, and groEL genes, (iii) admission IFA IgM titre of ≥1:12,800, (iv) 4-fold rise in IgM titre from paired samples. At least one of these criteria needs to be fulfilled for a positive scrub typhus diagnosis.

2, “***O. tsutsugamushi* isolation**”

Isolation of *O. tsutsugamushi* from cell culture

3, “**IFA**”

INDIRECT FLUORESCENCE ASSAY/INDIRECT FLUORESCENT ANTIBODY TEST/INDIRECT IMMUNOFLUORESCENCE ASSAY/INDIRECT IMMUNOFLUORESCENCE ANTIBODY TEST (IFA) detects antibody to scrub typhus in blood using immunofluorescent staining. The samples need to be assessed under fluorescence microscope. Often considered as a gold standard of scrub typhus diagnosis, despite the unideal sensitivity.

4, “**IIP**”

INDIRECT IMMUNOPEROXIDASE TEST detects antibody to scrub typhus in blood using an enzyme called peroxidase that will produce a change in colour. The samples can be assessed using light microscope.

5, “**PCR**”

POLYMERASE CHAIN REACTION (PCR)

The test is based on the detection of *O. tsutsugamushi* DNA sequences in patients' body fluid or tissues (e.g. eschar biopsy). PCR technique relies on amplification of DNA (Deoxyribonucleic acid) target sequences. The typical target are the 47kDa, 56 kDa, and groEL genes. The different PCR techniques for detecting *O. tsutsugamushi* DNA in patients' blood samples are: "Nested-PCR" (two step amplification often used for very low amount of DNA targets); “Real-Time PCR” or “Quantitative PCR” is based on the polymerase chain reaction technology, used to amplify and simultaneously quantify a targeted DNA molecule.

6, “**ELISA**”

ENZYME-LINKED IMMUNOSORBENT ASSAY (ELISA) is a method to detect antibody against scrub typhus in blood. It is less subjective than IFA. There are commercial test kits available.

7, “**ICT**”

IMMUNOCHROMATOGRAPHY TEST (ICT) is a method to detect antibody against scrub typhus in patient’s blood. It is relatively simpler to perform and results can be interpreted in about 15 minutes. Hence, it is often used as rapid point-of-care test.

8, “**Eschar presence**”

Diagnosis based on eschar presence, concordant history and clinical manifestation (including fever, other flu-like symptoms, rash, organ dysfunction). Eschar is necrotic skin lesion at the site of mite inoculation. It is a pathognomonic sign of scrub typhus.

9, “**Weil-Felix**”

Weil-Felix test is an agglutination test based on cross-reactions between antibodies against rickettsial infections with antigens of OX (OX 19, OX 2, and OXK) strains of Proteus species. Antibody against *O. tsutsugamushi* reacts with *P. mirabilis* OXK. It has low sensitivity and specificity.

10, “**History and clinical manifestation**”

Diagnosis based on history of travel to endemic area or foci of infection (“mite islands”) and clinical manifestation aside from eschar presence (including fever, other flu-like symptoms, rash, organ dysfunction).

11, “**Other**”

12, “**Unknown**”
